# Epigenome-wide association study of incident type 2 diabetes in Black and White participants from the Atherosclerosis Risk in Communities Study

**DOI:** 10.1101/2023.08.09.23293896

**Authors:** Sowmya Venkataraghavan, James S. Pankow, Eric Boerwinkle, Myriam Fornage, Elizabeth Selvin, Debashree Ray

**Affiliations:** Department of Epidemiology, Bloomberg School of Public Health, Johns Hopkins University, Baltimore, Maryland, United States of America; Division of Epidemiology and Community Health, School of Public Health, University of Minnesota, Minneapolis, Minnesota, United States of American; The UTHealth School of Public Health, Houston, Texas, United States of America; Brown Foundation Institute for Molecular Medicine, The University of Texas Health Science Center, Houston, Texas, United States of America; Welch Center for Prevention, Epidemiology, & Clinical Research, Johns Hopkins University, Baltimore, Maryland, United States of America; Department of Biostatistics, Bloomberg School of Public Health, Johns Hopkins University, Baltimore, Maryland, United States of America

## Abstract

DNA methylation studies of incident type 2 diabetes in US populations are limited, and to our knowledge none included individuals of African descent living in the US. We performed an epigenome-wide association analysis of blood-based methylation levels at CpG sites with incident type 2 diabetes using Cox regression in 2,091 Black and 1,029 White individuals from the Atherosclerosis Risk in Communities study. At an epigenome-wide significance threshold of 10^−7^, we detected 7 novel diabetes-associated CpG sites in *C1orf151* (cg05380846: HR= 0.89, *p* = 8.4 × 10^−12^), *ZNF2* (cg01585592: HR= 0.88, *p* = 1.6 × 10^−9^), *JPH3* (cg16696007: HR= 0.87, *p* = 7.8 × 10^−9^), *GPX6* (cg02793507: HR= 0.85, *p* = 2.7 × 10^−8^ and cg00647063: HR= 1.20, *p* = 2.5 × 10^−8^), chr17q25 (cg16865890: HR= 0.8, *p* = 6.9 × 10^−8^), and chr11p15 (cg13738793: HR= 1.11, *p* = 7.7 × 10^−8^). The CpG sites at *C1orf151*, *ZNF2, JPH3* and *GPX6*, were identified in Black adults, chr17q25 was identified in White adults, and chr11p15 was identified upon meta-analyzing the two groups. The CpG sites at *JPH3* and *GPX6* were likely associated with incident type 2 diabetes independent of BMI. All the CpG sites, except at *JPH3*, were likely consequences of elevated glucose at baseline. We additionally replicated known type 2 diabetes-associated CpG sites including cg19693031 at *TXNIP*, cg00574958 at *CPT1A*, cg16567056 at *PLBC2*, cg11024682 at *SREBF1*, cg08857797 at *VPS25*, and cg06500161 at *ABCG1*, 3 of which were replicated in Black adults at the epigenome-wide threshold. We observed modest increase in type 2 diabetes variance explained upon addition of the significantly associated CpG sites to a Cox model that included traditional type 2 diabetes risk factors and fasting glucose (increase from 26.2% to 30.5% in Black adults; increase from 36.9% to 39.4% in White adults). We examined if groups of proximal CpG sites were associated with incident type 2 diabetes using a gene-region specific and a gene-region agnostic differentially methylated region (DMR) analysis. Our DMR analyses revealed several clusters of significant CpG sites, including a DMR consisting of a previously discovered CpG site at *ADCY7* and promoter regions of *TP63* which were differentially methylated across all race groups. This study illustrates improved discovery of CpG sites/regions by leveraging both individual CpG site and DMR analyses in an unexplored population. Our findings include genes linked to diabetes in experimental studies (e.g., *GPX6*, *JPH3,* and *TP63*), and future gene-specific methylation studies could elucidate the link between genes, environment, and methylation in the pathogenesis of type 2 diabetes.

## INTRODUCTION

Diabetes is a chronic medical condition that is marked by elevated glycemia and can lead to major complications such as cardiovascular disease, peripheral artery disease, kidney disease, neuropathy, and retinopathy.^1^ There are major social disparities in diabetes; the incidence of diabetes is substantially higher among Black women (2.5 times) and men (1.4 times) compared to their White counterparts.^2^ Black adults with diabetes have an increased risk of developing retinopathy and kidney disease^3^ and are more likely to die from cardiovascular disease^4^ than White adults with diabetes. As of 2019, 14.7% of US adults had diabetes (both diagnosed and undiagnosed) of which 90-95% are expected to have type 2 diabetes.^5^ Type 2 diabetes is a complex disease with both genetic and environmental risk factors. While the earlier genome-wide association studies (GWAS) of diabetes exclusively studied participants of European ancestry, the recent ones are large-scale multi-ancestry studies detecting >250 loci in each study.^6, 7^ Some of the strongest type 2 diabetes-associated genetic variants reside at or near *PPARG*, *SLC30A8* and *TCF7L2*.^8^ Other risk factors for type 2 diabetes include age, obesity, sedentary lifestyle, and family history of diabetes.^9^

Environment and genetics can both change how regulatory processes influence gene expression without an underlying change in DNA sequence.^10^ These changes, referred to as epigenetic modifications, can contribute to changes in phenotype.^11^ The most widely studied epigenetic modification is DNA methylation (DNAm) where methyl groups are added to carbon position 5 on cytosine bases that are adjacent to guanine bases (CpG sites) across the genome.^10^ DNAm changes in response to lifetime environmental exposures may contribute to medical conditions in middle-aged and older adults.^11^ DNAm patterns may reflect social risk factors which may disproportionately affect Black individuals including exposure to racism,^12^ poverty,^13^ residential segregation, and air pollution^14^. Additionally, significant differences in DNAm levels at some CpG sites have been observed between Black and White infants at birth,^15^and a higher number of age-associated differentially methylated CpG sites have been found in Black adults compared to White adults,^16^ which could play a role in age-related diseases. DNAm is also partially genetically regulated with the average heritability of blood-based DNAm levels at CpG sites across the genome estimated to be 0.09 ± 0.02 (mean ± standard deviation) with >9% of CpG sites exhibiting heritability >0.3.^17^

Several epigenome-wide association studies (EWAS) examining association of DNAm levels with type 2 diabetes have identified and replicated CpG sites on or near several genes including *ABCG1*, *TXNIP*, *SREBF1* and *CPT1A*.^18–21^ EWAS of type 2 diabetes have predominantly been cross-sectional^19, 20, 22–24^, which cannot distinguish between DNAm changes that precede type 2 diabetes onset and DNAm changes that occur as a consequence of type 2 diabetes or diabetes medication. A few EWAS on incident type 2 diabetes exist; however, majority of these studies were conducted in European populations outside the US.^18, 21, 25^ In the US, a study of 1,312 American Indians from the Strong Heart Study identified several DNAm loci associated with HOMA-IR and fasting glucose that were also associated with incident type 2 diabetes.^26^

We sought to explore the epigenomic landscape of blood-based DNAm associated with incident type 2 diabetes in US middle-aged Black and White adults from the Atherosclerosis Risk in Communities (ARIC) study. We implemented epigenome-wide association analysis of DNAm levels at over 480,000 CpG sites, including examination of associations of previously discovered type 2 diabetes-associated CpG sites. We estimated the variation in incident type 2 diabetes accounted for by significantly associated CpG sites when added to traditional type 2 diabetes risk factors, and examined differentially methylated regions in individuals with incident type 2 diabetes versus those without.

## METHODS

### Study Participants

ARIC is a prospective cohort of adults aged 45-64 years at baseline from Forsyth County, North Carolina; Jackson, Mississippi; northwest suburbs of Minneapolis, Minnesota; and Washington County, Maryland. The baseline visit was between 1987-1989 with follow-up visits (Visits 2-9) occurring between 1990-present.^27^ The ARIC study research protocol was approved by the Institutional Review Board at each participating university. Written informed consent was obtained from participants including for genetic studies. Participants reported self-identified race at baseline from options “Black”, “White”, “Asian”, and “American or Alaskan Indian” in a questionnaire. For our analyses, we considered Black (n=2,796) and White participants (n=1,130) with DNAm data available from either Visit 2 or 3. We excluded participants from analyses if they did not have measured, imputed or estimated white blood cell (WBC) differentials (Black adults n=67, White adults n=0), had prevalent diabetes (classified based on self-reported doctor diagnosis or medication use) on or before the visit at which DNAm was measured (Black adults n=521, White adults n=65), had missing covariate measurements (Black adults n=66, White adults n=1), or had missing principal components (PCs) of ancestry computed using array-based SNP genotype data (Black adults n=51, White adults n=35). We had 2,091 Black and 1,029 White participants in our primary analysis. We conducted all our analyses stratified by race since exposures (DNAm levels) were measured separately by race group and a pooled analysis of all participants could be biased due to potential batch effects.

### Measurement of Outcome

We identified new diabetes cases based on self-reported doctor diagnosis or diabetes medication use assessed from the visit at which their DNAm measurements were taken (time origin) until 2019 either during an ARIC visit or through annual/semi-annual telephone calls.^28, 29^ Date of incident diabetes report was used as a proxy for date of diagnosis. Non-cases were censored at the time of loss to follow-up or administratively censored on 12/31/2019. Note, ARIC did not distinguish between type 1 and type 2 diabetes cases. We assume the vast majority are type 2 diabetes cases since in general 90-95% of diabetes instances are type 2 diabetes^5^, and type 1 diabetes diagnosis would have been rare in middle-aged people at this time (ARIC participants were >=45 years at baseline in 1987-1989).

### Measurement of DNAm Levels

DNA was extracted from peripheral blood leukocyte samples. DNAm levels at individual CpG sites were measured using the Illumina Infinium HumanMethylation450 BeadChip array also known as the HM450K array. Degree of methylation was determined using Illumina GenomeStudio 2011.1 Methylation module 1.9.0 software, and background correction was performed. DNAm levels at each CpG site, represented as beta (*β*) values with range 0-1 (0 is non-methylated and 1 is completely methylated), was estimated as the ratio of intensity of the methylated probe to the intensity of the methylated probe + unmethylated probe. Sample-level and CpG site-level quality control (QC) steps were undertaken. A total of 2,796 Black and 1,139 White participants were retained after sample-level QC. In Black adults, additional CpG site-level QC were applied. Following background correction and QC, Beta Mixture Quantile dilation (BMIQ) normalization was performed to reduce technical variation and biases of Infinium I and II probes. Additional details on DNAm measurement, quality control measures, and normalization are provided in **Supplementary Methods**. For our analyses, we examined CpG sites on autosomes only, which resulted in 470,161 and 469,973 CpG sites for Black and White adults respectively. All genomic coordinates are given in NCBI Build GRCh37/UCSC hg19.

### Measurement of Covariates

Since blood consists of multiple cell types with heterogenous DNAm profiles, we need to adjust for cell composition variability.^30^ A white blood cell (WBC) differential was measured in 175 Black participants during baseline visit. WBC proportions of neutrophils, lymphocytes, monocytes, eosinophils, and basophils were imputed in remaining Black participants using the measured subset as reference and the Houseman et. al imputation algorithm.^31, 32^ Cell type proportions in White participants were estimated using the estimateCellCounts function in minfi R package^33^ based on its in-built HM450K reference dataset only available for Europeans. Further details on cell type measurements are provided in **Supplementary Methods**. Weight was measured using a zeroed and calibrated scale.^34^ BMI was calculated as weight (in kilograms) divided by height (in meters) squared at time origin. Serum glucose (in mg/dL) was measured among participants fasting for 8 hours or more at time origin. Cigarette smoking status (current, former, never) and education level (less than high school, high school or equivalent, greater than high school) were assessed using questionnaires at time origin.^35^ Other covariates we included were age in years, self-reported sex coded as male/female, and genetic PCs. While both SNP-based PCs and methylation-based PCs can effectively account for population stratification arising from DNAm levels that vary by genetic ancestry, SNP-based PCs maximize power.^36^ We obtained SNP-based genetic PCs using PC-AiR^37^ function from R package GENESIS^38^ on existing genotype data from the Exome Chip array.

### Statistical Analyses

#### Adjustment of batch effects and cell type proportions

It is critical to adjust for potential batch effects when analyzing DNAm data. We used a linear mixed model for batch effect adjustment as had been done in previous studies using ARIC DNAm data.^39–41^ To do so, we analyzed DNAm data on 2,091 Black and 1,029 White adults separately in two stages. In stage 1, a linear regression model was fit with β-values of each CpG site as the dependent variable and adjusted for technical covariates and WBC proportions. Specifically, for Black adults, the technical covariates included chip ID, chip row, study center and visit, and the WBC proportions included the cell types neutrophils, lymphocytes, monocytes, and eosinophils. For White adults, the technical covariates included chip ID, chip row, study center, visit and project (unlike Black adults, DNAm data in White adults were measured as part of multiple projects), and the WBC proportions included cell types B, CD4^+^ T, CD8^+^ T, granulocytes, monocytes, and NK. We included chip ID as a random effect while chip row, study center, visit, project (when applicable) and WBC proportions were modeled as fixed effects. We obtained β-value residuals adjusted for batch effects and cell type proportions from this stage 1 model.

#### Time-to-event analysis

In stage 2, we conducted an epigenome-wide time-to-event analysis by fitting a Cox proportional hazards model with incident type 2 diabetes as outcome and the β-value residual of each CpG site obtained from stage 1 as exposure. We aligned the participants at time origin by the visit at which their blood samples for DNAm measurement were collected (visit 2 or 3). Our primary model (Model 1) for DNAm and incident type 2 diabetes association included covariate adjustments for age, sex, smoking status, education level, and the first 10 genetic PCs. For each covariate in the model, we checked if proportional hazards assumption was met for each race group by using Schoenfeld residual and cumulative martingale residual plots for continuous variables, and plots of log-log transformed survival and cumulative hazards against survival time for categorical variables.^42^ We meta-analyzed race-stratified results from our primary model using fixed-effects inverse-variance meta-analysis from the R package meta.^43^ We declared a CpG site as significantly associated with incident type 2 diabetes at a Bonferroni-corrected epigenome-wide significance threshold of 10^−7^(=0.05/480,407 CpG sites). We reported estimated hazard ratio (HR) for each significant CpG site, which represents change in the risk of developing type 2 diabetes per percent increase in DNAm β-values at the CpG site adjusted for model covariates.

We additionally fit secondary models to assess if significant CpG sites from Model 1 were associated with incident type 2 diabetes independent of BMI or fasting glucose. Model 2 included all covariates from Model 1 along with BMI as a continuous covariate and BMI x time interaction term in both race groups to model potential BMI-time dependence and ensure the proportional hazards assumption was met. Model 3 included all covariates from Model 1 along with time-varying fasting glucose effects that differed across periods of time since time origin. Details on the modeling choices of covariates in Models 2 and 3 are provided in **Supplementary Methods**. Thereafter, we meta-analyzed race-stratified results for each model to examine if the significant CpG sites remained statistically significant after BMI or fasting glucose adjustment and if there was any attenuation in log hazard ratios. For this, we calculated percent change in effect size as 100 × (log *HR*_no-BMI-adj_ − log *HR*_BMI-adj_)/ log *HR*_no-BMI-adj_ (similarly for fasting glucose), where a positive value is indicative of attenuation in effect size due to adjustment. A 0-3% change in effect size was considered minimal change.

#### Sensitivity analyses

We first assessed if removal of first-degree relatives qualitatively influenced our results since we did not remove related individuals in any of our analyses owing to low average heritability across genome-wide DNAm sites^17^. Second, we assessed if our findings are sensitive to different definitions of type 2 diabetes in ARIC. Third, we compared signals from the BMI-adjusted and the fasting glucose-adjusted secondary models with and without the proportional hazards assumption being met. Further details on these sensitivity analyses can be found in **Supplementary Methods**.

#### Estimation of variance explained

To estimate how much of the variation in incident type 2 diabetes is accounted for by the major non-genetic risk factors and the significant CpG sites identified in this study, we considered nested Cox proportional hazards models for Black and White adults separately. The first model “Cov” included the covariates age, sex, smoking status, education level and the first 10 genetic PCs. The second model “Cov + FG + BMI” included BMI and fasting glucose in addition to covariates in model “Cov”. To meet the proportional hazards assumption in the “Cov + FG + BMI” model, we included BMI, BMI x time interaction term, and time-varying effects of fasting glucose (i.e., different effect sizes across periods of time since time origin, as described under “**Time-to-event analysis**”). The third model “Cov + FG + BMI + DNAm” additionally included all the CpG sites significantly associated with incident type 2 diabetes in each race group in our primary model analysis. We determined variance of incident type 2 diabetes explained by the covariates in each model using Royston’s measure of explained variation for censored survival data (also known as Royston-D R^2^).^44^

#### Differentially methylated regions

We conducted differentially methylated region (DMR) analysis across two broadly defined categories. Gene-region specific DMRs were identified by analyzing grouped CpG sites within each gene region including 1500 bp ahead of transcription start site (TSS1500), 200 bp ahead of transcription start site (TSS200), 1^st^ exon of gene (Exon1), gene body regions post 1^st^ exon (Genebody), and 3’ untranslated region (3’UTR). The gene-region size distribution is depicted in **Supplementary Figure 1**. Gene-region agnostic DMRs were regions of CpG site associations in close proximity (within 1 to 500 bases) identified regardless of well-defined gene or gene-region boundaries. For both the gene-specific and gene-agnostic DMR analyses, we identified a region as DMR if it contained ≥ 3 CpG sites and had a Šidák-corrected p-value ≤ 0.05.^45, 46^ We marked regions as risk-increasing or risk-decreasing if all the estimated hazard ratios of individual CpG sites in the region were >1 or <1 respectively. A region was marked “mixed-effect” if it contained CpG sites where some had risk-increasing effects and others had risk-decreasing effects. Additionally for the gene-agnostic DMRs, we annotated them with any overlapping gene-regions obtained using the `fullannotInd` dataset based on Illumina methylation annotation. Specific details on gene-region specific and gene-region agnostic DMR analyses are provided in **Supplementary Methods**.

### Follow-up analyses

#### Candidate CpG site lookup

Apart from identifying potentially novel differentially methylated CpG sites in individuals with incident type 2 diabetes versus those without, we explored how many of the previously identified type 2 diabetes-associated CpG sites were replicated, particularly in Black adults since previous DNAm studies of incident type 2 diabetes did not consider individuals of African descent. We compiled a list of EWAS of both incident and prevalent type 2 diabetes from the NCBI PubMed database by searching the terms “DNA methylation”, “type 2 diabetes” and “epigenome-wide association analysis” or “EWAS” as of July 21, 2023. We included EWAS that used DNAm data measured through the Illumina platform (HM450K array or other variation of the Illumina Methylation array such as 27K, EPIC) as either exposure or outcome based on study design. We shortlisted a total of 17 studies including studies on DNAm from various tissues such as pancreatic islets, liver biopsy, subcutaneous and visceral adipose tissues, and whole blood (**Supplementary Table 1**).^18–26, 47–54^ Study-specific significance thresholds were used to obtain candidate CpG sites from each study. A total of 18,131 candidate CpG sites (unique candidate CpG sites: 419 from blood, 15,728 from adipose, 287 from liver, and 1,924 from pancreas) were looked up in the results from our primary time-to-event analysis.

#### Expression quantitative trait methylation (eQTM) lookup

We investigated if there was any evidence of association of the identified CpG sites with expression of the nearest genes in whole blood by querying our significantly associated CpG sites in the BBMRI-NL atlas.^55^ This atlas contains information on *cis*-eQTMs (association of gene expression with CpG sites within 250 Kb radius of transcription start site) identified at FDR < 5% in a sample of 3,841 Dutch participants from the BIOS consortium.^56^

## RESULTS

### Characteristics of study participants at time origin

Our analytic dataset included 2,091 Black and 1,029 White participants (**Table 1**). Due to the design of the ARIC Study, the majority (90%) of Black participants were enrolled at the Jackson, Mississippi Field Center. More than 60% of participants were female. DNAm was obtained from visit 2 blood samples for 87% of Black participants and 75% of White participants; the remaining participants had their DNAm measured in visit 3 samples. Black participants were on average younger, were less likely to have high school education, had higher BMI, and had higher fasting glucose levels than White participants.

**Table 1:** Study participant characteristics across race groups.

|  | **Black adults** | **White adults** | **All** |
| --- | --- | --- | --- |
|  | *n=2,091* | *n=1,029* | *n=3,120* |
| **Study center** |  |  |  |
| Forsyth County, North Carolina | 205 (9.8%) | 919 (89.3%) | 1123 (36.0%) |
| Jackson, Mississippi | 1886 (90.2%) | 0 (0%) | 1883 (60.4%) |
| Minneapolis, Minnesota | 0 (0%) | 87 (8.5%) | 87 (2.8%) |
| Washington County, Maryland | 0 (0%) | 23 (2.2%) | 23 (0.7%) |
| **Gender** |  |  |  |
| Female | 1314 (62.8%) | 604 (58.7%) | 1916 (61.5%) |
| Male | 777 (37.2%) | 425 (41.3%) | 1200 (38.5%) |
| **Visit** |  |  |  |
| Visit 2 | 1827 (87.4%) | 772 (75%) | 2596 (83.3%) |
| Visit 3 | 264 (12.6%) | 257 (25%) | 520 (16.7%) |
| **Age** |  |  |  |
| Mean (SD) | 56.7 (5.78) | 60.2 (5.4) | 57.9 (5.88) |
| **BMI** |  |  |  |
| Mean (SD) | 29.8 (6.22) | 26.1 (4.42) | 28.6 (5.94) |
| Missing | 4 (0.2%) | 0 (0%) | 4 (0.1%) |
| **Fasting glucose** |  |  |  |
| Mean (SD) | 106.4 (18.7) | 101.8 (13.14) | 104.7 (17.07) |
| Missing | 276 (13.2%) | 17 (1.7%) | 293 (9.4%) |
| **Smoking status** |  |  |  |
| Current smoking | 540 (25.8%) | 195 (19.0%) | 733 (23.5%) |
| Former smoking | 633 (30.3%) | 396 (38.5%) | 1028 (33.0%) |
| Never smoking | 918 (43.9%) | 438 (42.6%) | 1355 (43.5%) |
| **Education level** |  |  |  |
| No high school | 782 (37.4%) | 124 (12.1%) | 905 (29.0%) |
| High school | 592 (28.3%) | 445 (43.2%) | 1036 (33.2%) |
| More than high school | 717 (34.3%) | 460 (44.7%) | 1175 (37.7%) |

### Incident type 2 diabetes associations included 5 novel CpG sites from Black adults, 1 novel site each from White adults and all participants, and 6 previously discovered sites

The Manhattan plots (**Figure 1**) and the QQ plots (**Supplementary Figure 2**) across race groups do not indicate any remarkable inflation of p-values from our epigenome-wide association analysis. At an epigenome-wide significance threshold of 10^−7^, we identified 13 CpG site associations, most of which were identified in Black participants likely due to their higher sample size. The Volcano plots (**Figure 1**) highlight the wide range of effect sizes of the associated CpG sites.

**Figure 1.**
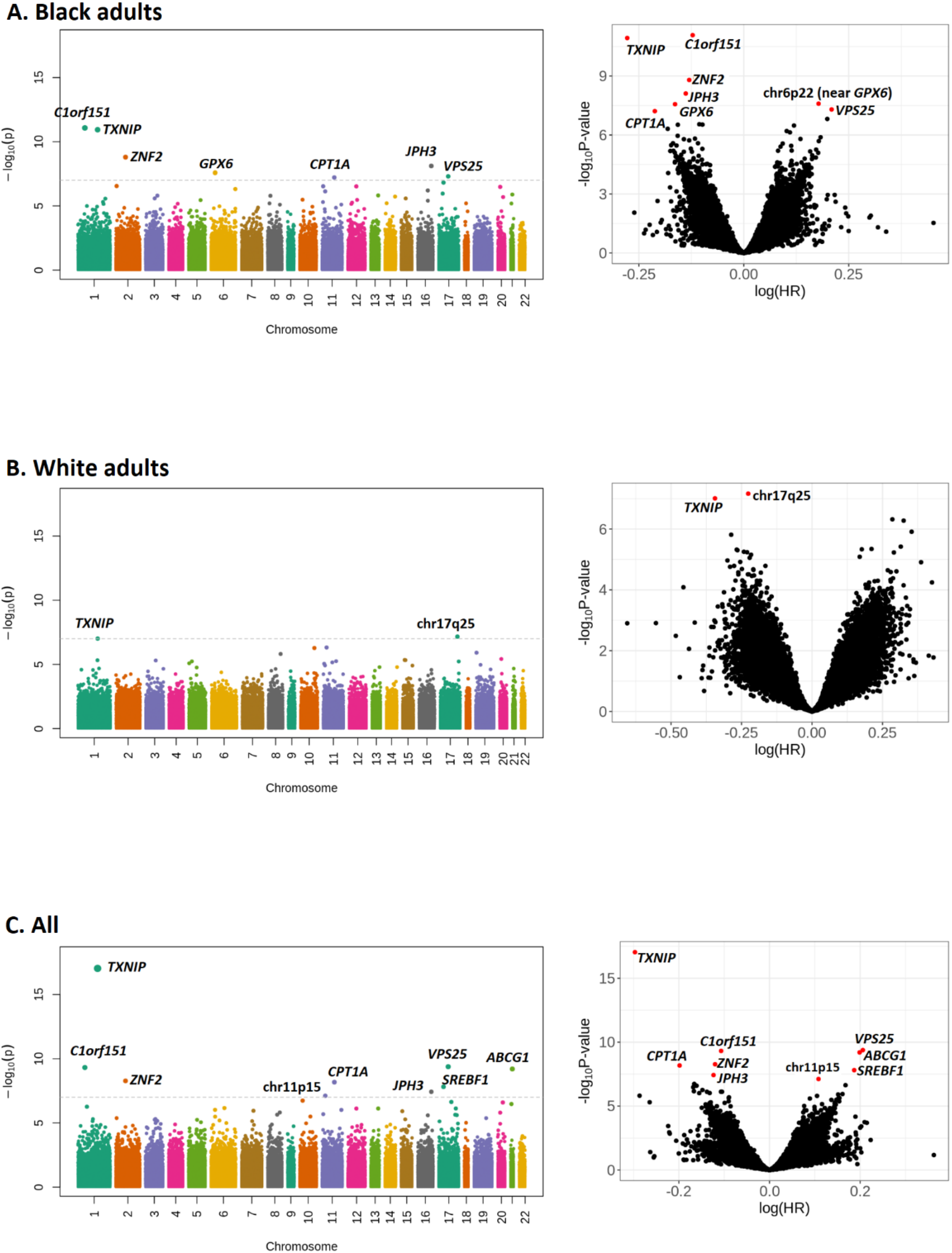
Manhattan and volcano plots of epigenome-wide CpG site association of incident type 2 diabetes using Cox proportional hazards model across race groups and the combined meta-analyzed group. The model was adjusted for age, sex, smoking status, education level, and the first 10 genetic principal components. Technical covariates (chip ID, chip row, study center, visit, project) and cell type proportions were adjusted too. The significant CpG sites are annotated by the nearest gene or the locus if it is a gene-free region.

Two novel CpG sites located on or near *GPX6* at chr6p22 were associated with incident type 2 diabetes in Black adults alone (**Supplementary Figure 3**). cg02793507 in the gene body of *GPX6* was associated with a decreased risk of developing type 2 diabetes (HR= 0.85, *p* = 2.7 × 10^−8^) while cg00647063 near *GPX6* was associated with an increased risk per percent increase in DNAm β values after adjustment of model covariates (HR= 1.20, *p* = 2.5 × 10^−8^) (**Table 2**). Three other novel associations include CpG sites in the 3’-UTRs of *C1orf151* (HR= 0.89, *p* = 8.4 × 10^−12^), *ZNF2* (HR= 0.88, *p* = 1.6 × 10^−9^), and *JPH3* (HR= 0.87, *p* = 7.8 × 10^−9^) (**Supplementary Figures 4-5** and **Figure 2**). Further, we replicated 3 CpG sites previously discovered for incident or prevalent type 2 diabetes at chr1q21 in the 3’-UTR of *TXNIP* (HR= 0.76, *p* = 1.2 × 10^−11^), chr11q13 in the 5’-UTR of *CPT1A* (HR= 0.81, *p* = 6.1 × 10^−8^) and chr17q21 in the gene body region of *VPS25* (HR= 1.23, *p* = 5 × 10^−8^) (**Supplementary Figures 6-8**). When we restricted our analysis to an unrelated subset of Black participants, cg02793507 at *GPX6* did not remain significant (HR= 0.85, *p* = 1.1 × 10^−6^) although there was no remarkable change in effect size or its 95% confidence interval (**Supplementary Table 2**). In fact, our sensitivity analysis with and without removal of first-degree relatives did not reveal any qualitative difference in hazard ratio estimates of top 50 CpG sites across race groups (**Supplementary Figure 9**). When using a more stringent definition for type 2 diabetes cases (self-reported medication use only), both cg02793507 (HR= 0.91, *p* = 0.002) and cg00647063 (HR= 1.1, *p* = 1.1 × 10^−7^) at *GPX6* were no longer significant (**Supplementary Table 3**).

**Figure 2:**
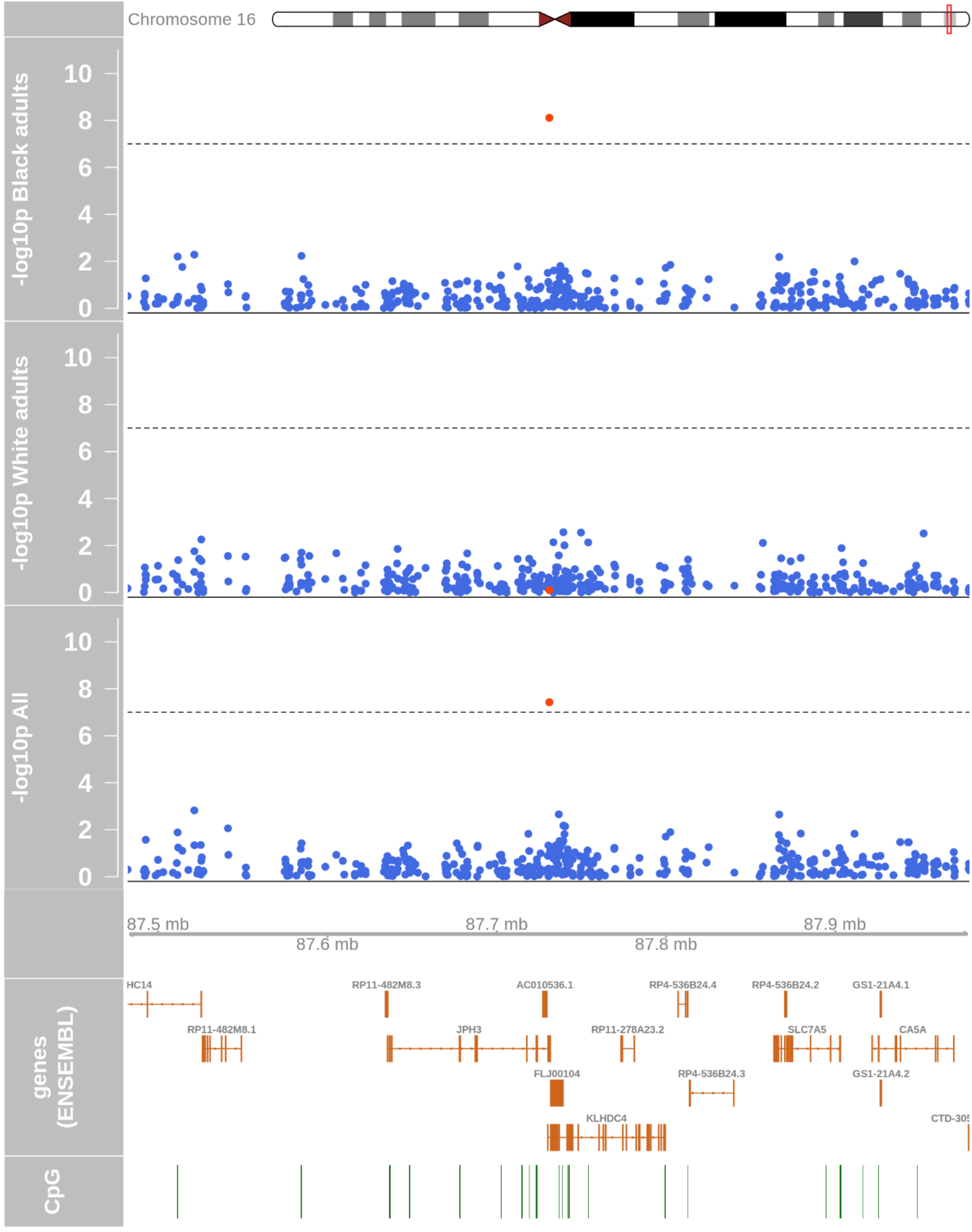
Regional association plot of CpG sites at chr16q24 near *JPH3* in an epigenome-wide incident type 2 diabetes analysis across race groups. A Cox proportional hazards model adjusting for age, sex, smoking status, education level, and the first 10 genetic principal components was fit. Technical covariates (chip ID, chip row, study center, visit, project) and cell type proportions were adjusted too. Negative log-transformed p-values of association are plotted against chromosome coordinates, genes and CpG islands. Black dashed lines correspond to Bonferroni-corrected epigenome-wide significance threshold of 10^−7^.

**Table 2:** Significantly associated CpG sites with incident type 2 diabetes at p < 10^−7^ in one or more race groups for the primary model analysis wherein a Cox proportional hazards model adjusted for age, sex, smoking status, education level and first 10 genetic PCs was fit. Technical covariates (chip ID, chip row, study center, visit, project) and cell type proportions were adjusted too. Entries are **bold-faced** if the CpG site is significantly associated with incident type 2 diabetes in that race group. “PH test p-value” represents the p-value of Schoenfeld residuals test used to test the proportional hazards assumption. “eQTM target gene p-value” indicates association between DNAm levels and target gene (eQTM gene) in the BBMRI-NL atlas.

| Chr | Position (hg19) | CpG | Gene | Gene group | eQTM gene | eQTM target gene p-value | Black adults |  |  | White adults |  |  | All |  |
| --- | --- | --- | --- | --- | --- | --- | --- | --- | --- | --- | --- | --- | --- | --- |
|  |  |  |  |  |  |  | HR (95% CI) | P-value | PH test p-value | HR (95% CI) | P-value | PH test p-value | HR (95% CI) | P-value |
| chr1p36 | 19953389 | cg05380846* | *C1orf151* | 3'UTR |  | - | **0.89(0.85,0.92)** | **8.39E-12** | **0.120** | 1.08(0.96,1.22) | 0.22 | 0.18 | **0.9(0.87,0.93)** | **4.84E-10** |
| chr1q21 | 145441552 | cg19693031 | *TXNIP* | 3'UTR | *TXNIP* | 7.14E-08 | **0.76(0.7,0.82)** | **1.17E-11** | **0.001** | **0.71(0.62,0.8)** | **9.79E-08** | 0.10 | **0.74(0.69,0.8)** | **9.15E-18** |
| chr2q11 | 95847957 | cg01585592* | *ZNF2* | 3'UTR |  | - | **0.88(0.84,0.92)** | **1.59E-09** | **0.552** | 0.98(0.86,1.13) | 0.80 | 0.09 | **0.89(0.85,0.92)** | **5.33E-09** |
| chr6p22 | 28478052 | cg02793507* | *GPX6* | Body |  | - | **0.85(0.8,0.9)** | **2.71E-08** | **0.313** | 1.03(0.9,1.17) | 0.66 | 0.05 | 0.88(0.83,0.92) | 9.58E-07 |
| chr6p22 | 28510682 | cg00647063* | - | - |  | - | **1.2(1.12,1.27)** | **2.54E-08** | **0.414** | 0.92(0.8,1.06) | 0.26 | 0.10 | 1.15(1.08,1.21) | 3.26E-06 |
| chr11p15 | 6337206 | cg13738793* | - | - |  | - | 1.12(1.07,1.17) | 7.36E-07 | 0.293 | 1.11(1.01,1.21) | 0.04 | 0.16 | **1.11(1.07,1.16)** | **7.64E-08** |
| chr11q13 | 68607622 | cg00574958 | *CPT1A* | 5'UTR | *CPT1A* | 3.05E-20 | **0.81(0.75,0.87)** | **6.12E-08** | **0.156** | 0.86(0.75,0.98) | 0.03 | 0.15 | **0.82(0.77,0.88)** | **6.67E-09** |
| chr15q15 | 40599985 | cg16567056 | *PLCB2* | 1stExon; 5'UTR |  | - | 1.1(1.06,1.14) | 2.44E-07 | 0.133 | 1.08(0.98,1.2) | 0.12 | 0.17 | **1.1(1.06,1.14)** | **7.55E-08** |
| chr16q24 | 87731080 | cg16696007* | *JPH3* | 3'UTR |  | - | **0.87(0.83,0.91)** | **7.76E-09** | **0.382** | 0.98(0.87,1.12) | 0.79 | 0.17 | **0.88(0.85,0.92)** | **3.75E-08** |
| chr17p11 | 17730094 | cg11024682 | *SREBF1* | Body | *SREBF1* | 4.50E-15 | 1.22(1.13,1.31) | 1.53E-07 | 0.083 | 1.16(1.02,1.32) | 0.03 | 0.06 | **1.21(1.13,1.29)** | **1.54E-08** |
| chr17q21 | 40927699 | cg08857797 | *VPS25* | Body |  | - | **1.23(1.14,1.33)** | **5.00E-08** | **0.188** | 1.22(1.07,1.38) | 2.23E-03 | 0.22 | **1.23(1.15,1.31)** | **4.16E-10** |
| chr17q25 | 77669201 | cg16865890* | - | - |  | - | 0.94(0.89,0.99) | 0.01 | 0.316 | **0.8(0.73,0.87)** | **6.88E-08** | 0.23 | 0.9(0.86,0.94) | 7.52E-07 |
| chr21q22 | 43656587 | cg06500161 | *ABCG1* | Body | *ABCG1* | 2.22E-37 | 1.2(1.12,1.29) | 1.30E-06 | 0.114 | 1.27(1.13,1.43) | 8.62E-05 | 0.20 | **1.22(1.15,1.3)** | **6.30E-10** |

\*novel CpG site association with diabetes

We found 1 novel CpG site cg16865890 in a gene free region at chr17q25 (HR= 0.8, *p* = 6.9 × 10^−8^) in White adults alone (**Supplementary Figure 10**). This site was no longer significant in White participants when first degree relatives were removed from analysis (HR= 0.81, *p* = 1.6 × 10^−6^) or when a more stringent definition of type 2 diabetes was used (HR= 0.87, *p* = 4.0 × 10^−3^). We replicated the CpG site in the 3’-UTR of *TXNIP* (HR= 0.71, *p* = 9.8 × 10^−8^) in White adults too.

The meta-analysis of race-stratified results additionally identified 1 novel and 3 previously identified CpG sites, all of which were associated with increased risk of type 2 diabetes. The novel CpG site was in a gene-free region at chr11p15 (HR= 1.11, *p* = 7.7 × 10^−8^) (**Supplementary Figure 11**), which lost its epigenome-wide significance when a more stringent definition of type 2 diabetes was used (HR= 1.10, *p* = 5.4 × 10^−5^). The 3 type 2 diabetes-associated CpG sites we replicated include cg16567056, cg11024682 and cg06500161 and on the gene body regions of *PLCB2* (HR=1.09, *p* = 7.6 × 10^−8^), *SREBF1* (HR= 1.21, *p* = 1.5 × 10^−8^) and *ABCG1* (HR= 1.22, *p* = 6.3 × 10^−10^) respectively (**Supplementary Figures 12-14**). The latter 2 CpG sites were eQTMs for *SREBF1* (*p* = 4.5 × 10^−15^) and *ABCG1* (*p* = 2.2 × 10^−37^) respectively.

### CpG sites at *GPX6* and *JPH3* were likely associated with incident type 2 diabetes independent of BMI

Three of the novel CpG sites discovered in Black adults continued to be epigenome-wide significant after BMI adjustment: *C1orf151* (HR= 0.90, *p* = 1.8 × 10^−8^), *GPX6* (HR= 0.85, *p* = 6.4 × 10^−8^) and *JPH3* (HR= 0.87, *p* = 3.4 × 10^−9^) (**Supplementary Table 4**). There was 2% attenuation in effect size for the CpG site at *GPX6* and 1% increase in effect size at *JPH3* after BMI adjustment. Interestingly, we also found a BMI-dependent (cg00647063) CpG site association in *GPX6*, 32 Kb downstream of the BMI-independent site (cg02793507). The novel CpG site identified in White adults at chr17q25 was also significant after BMI adjustment (HR= 0.78, *p* = 8.3 × 10^−9^) with 1% increase in effect size. The well-known type 2 diabetes-associated CpG site on *TXNIP* remained significantly associated with incident type 2 diabetes after BMI adjustment (HR= 0.76, *p* = 2.2 × 10^−11^) with 1% attenuation in its effect size while the CpG sites at *ABCG1* and *CPT1A* were not, replicating the findings of a previous study on incident type 2 diabetes^21^. CpG sites identified in BMI-adjusted analysis across race-stratified and meta-analyzed groups were identical irrespective of whether BMI-time dependence was modeled to meet proportional hazards assumption or not (**Supplementary Figure 15**).

### Except *JPH3*, all discovered CpG sites were likely consequences of elevated blood glucose levels

cg16696007 in *JPH3* was the only CpG site that remained significant after fasting glucose adjustment with minimal effect size attenuation in both Black adults (HR= 0.87, *p* = 2.7 × 10^−9^) and the meta-analyzed group (HR= 0.88, *p* = 3.2 × 10^−9^). No CpG site we identified from White adults retained statistical significance after fasting glucose adjustment. We found fasting glucose-adjusted analysis in White adults was sensitive to whether proportional hazards assumption was met or not (**Supplementary Figure 16**).

### Modest increase in incident type 2 diabetes variance explained by DNAm levels at epigenome-wide significant CpG sites

In Black adults, variance explained by model "Cov" (age, sex, smoking status, education level and genetic PCs) was 1.7% while addition of BMI and fasting glucose to the model explained 26.2% of the variation (**Supplementary Figure 17**). In White adults, variance explained by models “Cov” and “Cov + FG + BMI” were 8.8% and 36.9% respectively. When DNAm levels of epigenome-wide significant CpG sites from each race group were further added, the explained variation increased to 30.5% in Black adults and 39.4% in White adults. We should interpret this improvement with caution since these estimates could be inflated due to the use of same study participants for discovery and explained variation calculation.

### CpG sites previously discovered from blood and adipose tissues were among our epigenome-wide significant CpG sites

Among the blood-based candidate CpG sites, the top associated CpG site was cg19693031 of *TXNIP* that was epigenome-wide significant in all race groups (**Supplementary Table 5**). The top associated adipose-based candidate CpG site was cg16567056 of *PLCB2* in Black adults and the meta-analyzed group, and cg17582466 in a gene free region at chr10q26 in White adults (HR= 1.38, *p* = 5.3 × 10^−7^) (**Supplementary Table 6)**. The top associated liver-based candidate CpG site was cg06533700 in a gene free region at chr1p22 in Black adults (HR= 0.88, *p* = 5.8 × 10^−4^) and the meta-analyzed group (HR= 0.88, *p* = 3.3 × 10^−5^), and cg24655262 at chr17p13 overlapping 3’-UTR and 1^st^ exon region of *C17orf59* in White adults (HR= 0.84, *p* = 1.9 × 10^−3^) (**Supplementary Table 7**). The top associated pancreas-based candidate CpG site was cg06690548 at chr4q31 on the gene body region of *SLC7A11* in Black adults (HR= 0.88, *p* = 2.3 × 10^−4^) and the meta-analyzed group (HR= 0.89, *p* = 6.4 × 10^−5^), and cg16434331 at chr17q21 on the gene body region of *SLC39A11* in White adults (HR= 1.32, *p* = 6.0 × 10^−4^) (**Supplementary Table 8**).

### Gene-regions annotated to 483 distinct genes were identified across race groups in gene-region specific DMR analysis

A total of 205 genes in Black adults and 151 genes in White adults had one or more gene-regions with risk differences for developing type 2 diabetes (**Supplementary Tables 9-10**). The meta-analyzed group additionally yielded 142 genes with one or more gene-region DMRs (**Supplementary Table 11**). Across race groups, the majority of gene-region DMRs were identified in the gene body regions post 1^st^ exon (**Figure 3**) that influenced risk in both directions likely because several of these Genebody regions were longer in length than other gene-regions (**Supplementary Figure 1**). More than 80% of differentially methylated gene-regions did not demonstrate a consistent direction of hazard ratio for individual CpG sites within that region. Black adults had the highest proportion (8%) of risk-decreasing DMRs of which the largest proportion (44%) was attributed to Genebody region. White adults had the highest proportion (10%) of risk-increasing DMRs of which the largest proportion (44%) belonged to TSS200 region.

**Figure 3:**
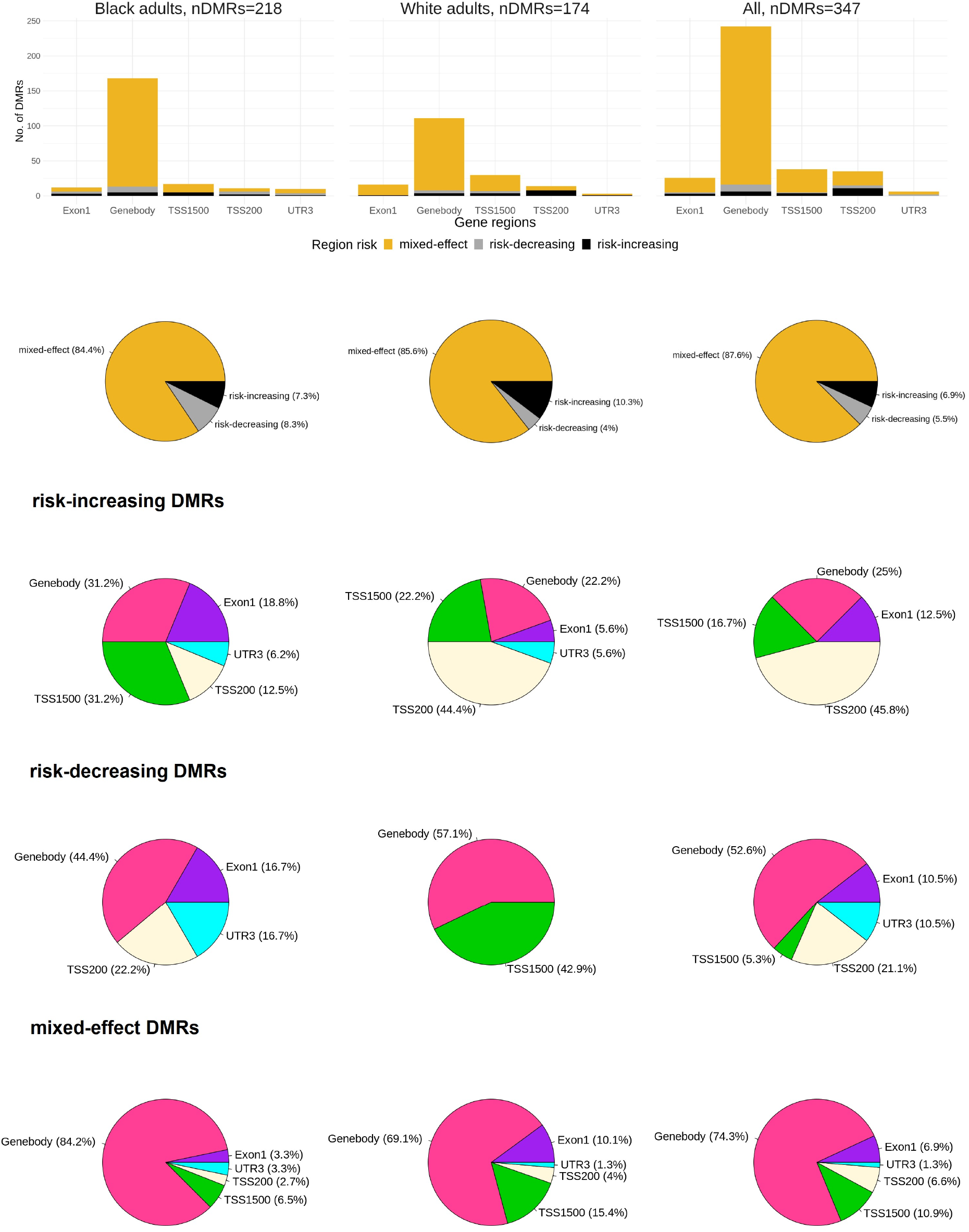
Distribution of risk-increasing, risk-decreasing and mixed-effect differentially methylated regions (DMRs) across gene regions in Black adults, White adults, and the meta-analyzed group in gene-region specific DMR analysis. ‘Mixed-effect’ DMR represents a region where some CpG sites have risk-increasing effect while others have risk-decreasing effect on incident type 2 diabetes.

### Gene-regions of *TP63* and *PCDHγ* were consistently detected in both race groups

In Black adults, gene regions TSS1500 (*p* = 8.3 × 10^−4^, n=11 CpG sites) and TSS200 (*p* = 2.6 × 10^−3^, n=9 CpG sites) of *TP63* were differentially methylated (**Figure 4**). Both of these *TP63* regions (TSS1500 *p* = 2.1 × 10^−5^, TSS200 *p* = 1.3 × 10^−3^) were similarly differentially methylated in the meta-analyzed group while only the TSS1500 region (*p* = 5 × 10^−3^) was differentially methylated in White adults. We also identified DMRs among the overlapping Genebody regions of the *PCDHγ* family of genes in both Black and White participants (**Supplementary Table 12**). The Genebody region of known type 2 diabetes gene *IGF2BP2* was significantly differentially methylated (*p* = 5.5 × 10^−3^, n=30 CpG sites) in the meta-analyzed group (**Supplementary Figure 18**). While Genebody region of *ANK2* was differentially methylated (*p* = 6.4 × 10^−4^, n=40 CpG sites) in Black adults (**Supplementary Figure 19**), the TSS1500 (*p* = 4.5 × 10^−3^, n=31 CpG sites), TSS200 (*p* = 0.015, n=28 CpG sites), Exon1 (*p* = 8.5 × 10^−3^, n=27 CpG sites) and Genebody (*p* = 0.024, n=30 CpG sites) regions of *ANK3* were differentially methylated in White adults (**Supplementary Figure 20**). All of the above DMRs consisted of some CpG sites that increased risk of developing type 2 diabetes while others decreased risk.

**Figure 4:**
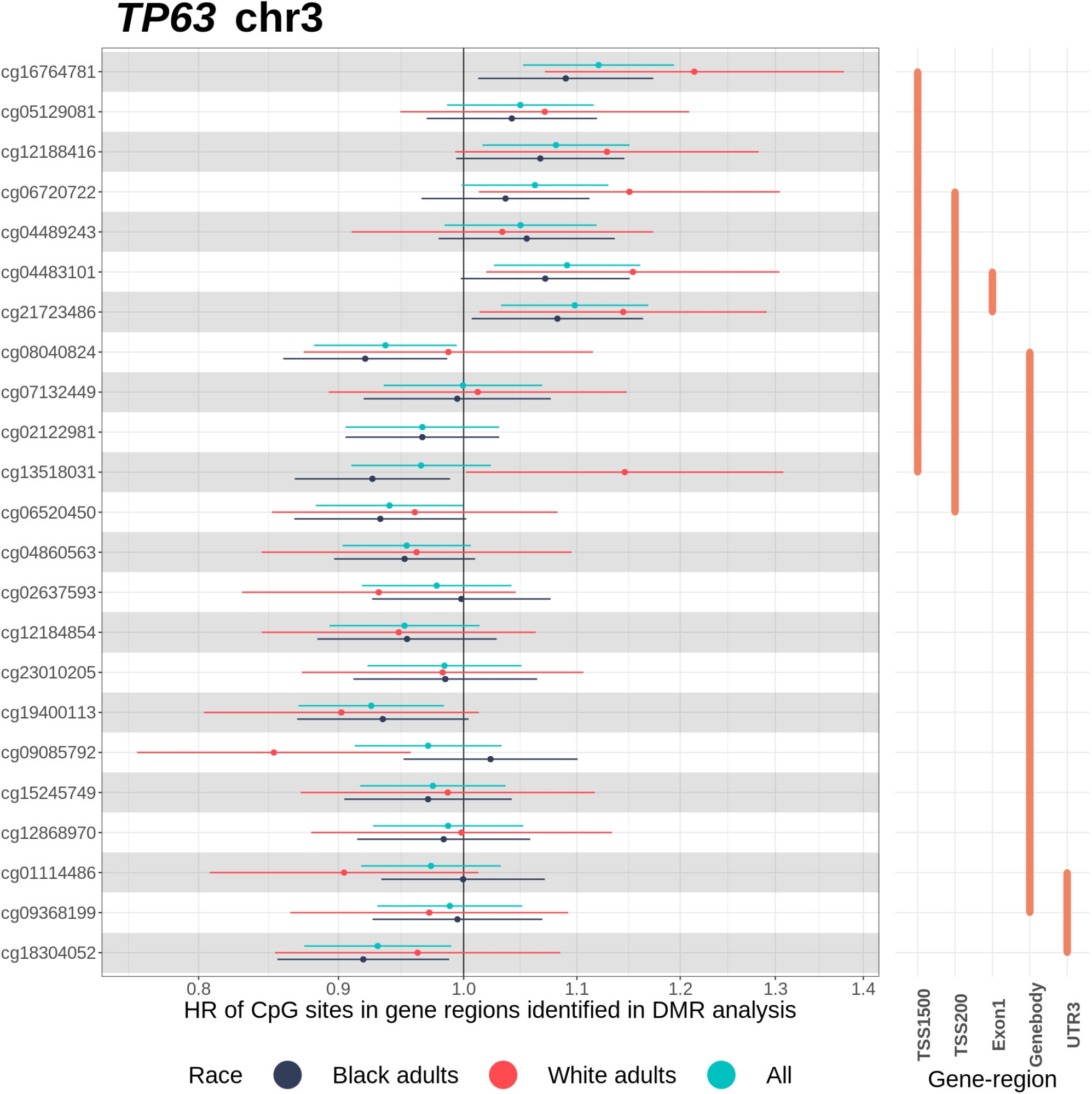
Effect size plot of CpG sites annotated to gene-regions of *TP63* on chromosome 3. The graph on the left depicts hazard ratio estimates of CpG sites with 95% confidence intervals across race groups obtained from Cox proportional hazards model adjusting for age, sex, smoking status, education level, and the first 10 genetic principal components. Technical covariates (chip ID, chip row, study center, visit, project) and cell type proportions were adjusted too. CpG sites are arranged from top to bottom in ascending order of chromosomal position. The graph on the right indicates the gene-region DMR groups that each CpG site was included in.

### Additional DMRs common to both race groups were identified from gene-region agnostic DMR analysis

We detected 85 DMRs in Black adults, 83 in White adults, and 99 in the meta-analyzed group (**Supplementary Table 13**). The majority of gene-region agnostic DMRs was risk-increasing across race groups (**Figure 5**). Black adults had the highest proportion of risk-decreasing DMRs, which mostly overlapped with exons. Across race groups, the distribution of risk-increasing and mixed-effect regions were largely similar, and a majority of these regions overlapped with TSS1500, TSS200 and Exon1 or promoter regions. We found 2 DMRs common across all race groups at the Šidák-corrected significance threshold of 5%: a risk-decreasing region on chr 5 comprising 7 CpG sites overlapping with *TMEM232* (Black adults *p* = 2.9 × 10^−2^, White adults *p* = 0.027, All *p* = 3.5 × 10^−10^) (**Supplementary Figure 21A**), and a risk-increasing region on chr 16 comprising 5 CpG sites overlapping with *ADCY7* (Black adults *p* = 1.5 × 10^−4^, White adults *p* = 0.011, All *p* = 1.7 × 10^−9^) (**Supplementary Figure 21B**). We found DMRs overlapping with *HLA-DPB1* and *HLA-DPA1* in both Black and White adults. In particular, for Black adults, 2 risk-increasing DMRs on chr 6 comprising 12 CpG sites (*p* = 4.8 × 10^−5^) and 9 CpG sites (*p* = 1.4 × 10^−10^) overlapped with *HLA-DPB1* and *HLA-DPA1* (**Supplementary Figures 21C-D**). On the other hand, in White adults, 1 risk-increasing DMR comprising 25 CpG sites (*p* = 6 × 10^−9^) overlapped with *HLA-DPB1* (**Supplementary Figure 21E**). A completely overlapping risk-increasing DMR between Black and White adults was near *SLC16A3*, where the DMR in Black adults (*p* = 2.7 × 10^−7^, n=5 CpG sites) was 335 bp longer than the DMR in White adults (*p* = 0.03, n=4 CpG sites) (**Supplementary Figures 21F-G**). Several DMRs identified in the meta-analyzed group included regions that had some overlap with DMRs identified in either Black or White adults.

**Figure 5:**
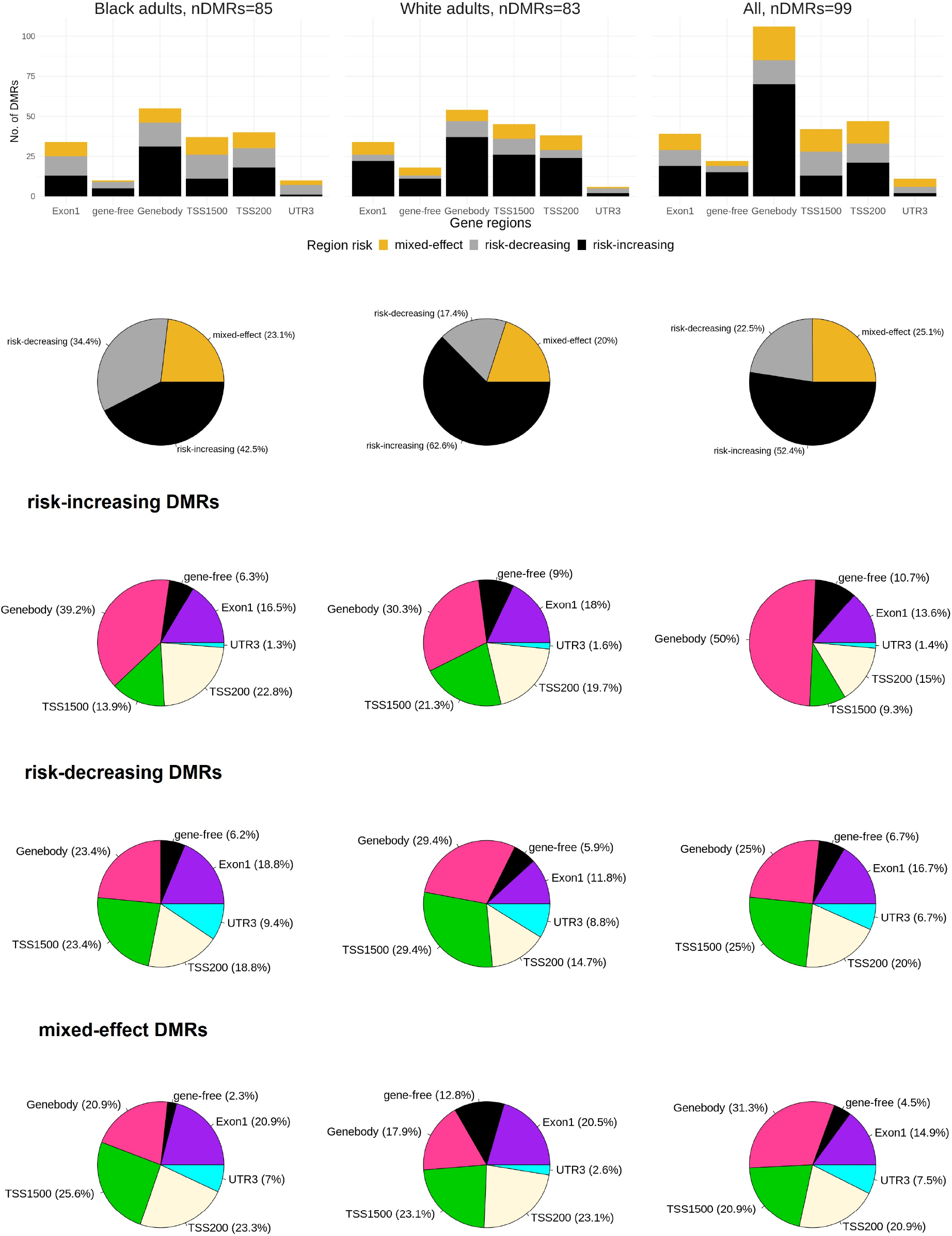
Distribution of risk-increasing, risk-decreasing and mixed-effect DMRs across gene-regions in Black adults, White adults, and the meta-analyzed group in the gene-region agnostic DMR analysis. ‘Mixed-effect’ DMR represents a region where some CpG sites have risk-increasing effect while others have risk-decreasing effect on incident type 2 diabetes.

## DISCUSSION

To our knowledge, this is the first prospective analysis of epigenome-wide DNA methylation and incident type 2 diabetes in a study that included individuals of African descent. Previous studies of incident type 2 diabetes have relied heavily on European and Indian Asian cohorts.^18, 21, 25, 53^ We discovered 7 novel CpG sites at epigenome-wide significance threshold and several differentially methylated regions at Šidák-corrected significance threshold for incident type 2 diabetes during a median follow-up of 17 years. These novel CpG sites were discovered primarily in Black adults. Race-specific findings from these analyses do not necessarily indicate inherent biological differences between race groups. Rather, these differences can arise due to differences in statistical power (e.g., from different sample sizes for each race group), differences in environmental stressors leading to downstream changes in DNAm levels (e.g., systemic racism, economic disparities, and other social factors that influence health), and differences in allele frequencies of variants upstream to DNAm levels since these race groups are enriched for different genetic ancestries^57^.

In Black adults, the CpG sites discovered on the gene body of *GPX6* and the 3’-UTR of *JPH3* were associated with decreased risk of type 2 diabetes, and had <3% attenuation in log hazard ratios when adjusted for BMI. CpG sites on gene body may be involved in splicing or increase in expression of the gene on which CpG site is located.^58, 59^ *GPX6* is a member of the Glutathione peroxidase family (GPx), involved in protection of cells from oxidative damage. GPx enzyme is a key indicator of oxidative stress^60^, and markers of oxidative stress have been linked to insulin resistance.^61^ Previous studies have identified associations of variants near *GPX6* with HbA1c,^62^ BMI-adjusted waist circumference^63^ and lipids^64^, and that circulating GPx protein levels were suppressed in individuals with diabetes compared to those without diabetes^60^. It is possible that methylation at the *GPX6* sites are simply surrogate markers for elevated glucose, rather than play a causal role in diabetes pathogenesis, because these associations were considerably attenuated upon adjustment for fasting glucose. The *JPH3* site, on the other hand, had <1% effect attenuation with fasting glucose adjustment. *JPH3* is expressed in human and mouse pancreatic beta cells; silencing of *JPH3* expression in mice reduced insulin secretion in response to glucose^65^; and there is a very strong genetic support for the involvement of common variants in *JPH3* on HbA1c^66^. Blood glucose levels typically increase steadily with age and may increase more steeply in individuals with diabetes several years before they are diagnosed^67^. The *JPH3* site could be reflective of early pathophysiological changes preceding type 2 diabetes. A neighboring CpG site in the gene body of *JPH3* in blood was found associated with pollution from exposure to heavy vehicles.^68^ Methylation in the promoter region of *JPH3* in sputum was associated with chronic mucous hypersecretion in former smokers.^69^ Future studies of methylation at *JPH3* could shed more light on the effects of environmental exposures, such as smoke exposure, on type 2 diabetes. Another novel CpG site in Black adults that decreased risk of type 2 diabetes was in the 3’-UTR of *ZNF2*. *ZNF2* may be involved in transcriptional regulation, and there is a strong genetic support for the involvement of rare variants in *ZNF2* on the glycemic trait fasting C-peptide.^70^

We found CpG sites identified in this study explained modest additional variance in type 2 diabetes risk beyond traditional risk factors and fasting glucose (a strong predictive biomarker) in both Black and White adults. The value of identifying type 2 diabetes-associated CpG site association lies in understanding biological pathways of type 2 diabetes. We predominantly discovered CpG sites that are not near genetic variants associated with type 2 diabetes risk. Genes near known type 2 diabetes-associated CpG sites such as *CPT1A* and *TXNIP* have, to our knowledge, not been implicated in GWAS of type 2 diabetes. Genes discovered in DNAm-type 2 diabetes association studies but not in GWAS of type 2 diabetes could point to changes in gene expression associated with type 2 diabetes predominantly due to epigenetics in the presence of weak or no effect of variation in genotype.

In our gene-region specific DMR analysis, we identified DMRs near the promoter region of *TP63* consistently across race-groups. TAp63, an isoform of *TP63*^71^, knockout mice were found to develop insulin resistance and glucose tolerance.^72^ Mice lacking TAp63 exhibited increased gluconeogenesis and over-expression of TAp63 increased insulin sensitivity.^73^ We identified a DMR in the gene body of *ANK2* in Black adults and 4 gene-region associations of *ANK3* in White adults. Mice homozygous for *ANK2* variants were found to have early-onset pancreatic beta cell dysfunction and increased insulin resistance.^74^ *ANK1*, another member of the same adapter protein family of ankyrins as *ANK2* and *ANK3*, is a known type 2 diabetes-associated gene.^6, 7^ A large proportion of the gene-region agnostic DMRs we identified were risk-increasing as opposed to predominantly mixed-effect DMRs identified in gene-specific DMR analysis. We did not detect any DMR overlapping with *GPX6* despite finding two CpG site associations near this gene; this limitation could be due to pre-selected regions not extending long enough to encompass both sites (**Supplementary Figure 22**). We note the value of performing both DMR and individual CpG site analysis: while analysis of CpG sites one at a time would uncover those with strong effects on type 2 diabetes, DMR analysis may highlight additional DNAm regions by leveraging dependence between DNAm levels of proximal CpG sites with weak effects.

Multiple CpG sites previously discovered in studies of prevalent and/or incident type 2 diabetes were replicated in this study at epigenome-wide significance. cg19693031 at *TXNIP*, was previously found to be associated with incident type 2 diabetes in Europeans and Indian Asians,^18, 21^ and prevalent type 2 diabetes in sub-Saharan Africans^20^. Similar to previous studies, cg19693031 at *TXNIP* showed the strongest association in our study.^18, 23, 25^ cg00574958 at *CPT1A* was found associated with incident type 2 diabetes in Europeans^25, 53^ and prevalent type 2 diabetes in sub-Saharan Africans^20^. cg08857797 at *VPS25* was suggestively associated (*p* < 10^−5^) with prevalent type 2 diabetes in a European meta-analysis.^53^ These blood-based candidates were epigenome-wide significant in Black adults. In our meta-analysis of Black and White adults, we replicated cg11024682 at *SREBF1* and cg06500161 at *ABCG1* discovered predominantly in populations outside the US^26, 75^. Through our gene-region agnostic DMR analysis, we replicated a CpG region near *ADCY7*. A CpG site cg02879453 (*p* = 3.5 × 10^−5^ in the meta-analyzed group) in the promoter region of *ADCY7* was previously implicated in a meta-analysis of incident type 2 diabetes in European populations.^21^ The directions of effects for all replicated findings were consistent with previous discovery.

Our study has several limitations. A key attribute of our study is the use of DNAm measurements from whole blood. While diabetes-relevant tissues such as pancreas and adipose may carry DNAm signatures different from whole blood, they are more patient invasive. This limits the functional insights we can gather on type 2 diabetes since DNAm levels vary across cell types and tissues. Blood sample collection is, however, less invasive and is done routinely. DNAm measurements are correlated across tissues for certain genes and CpG regions^76^ that can lead to valuable findings using blood-based DNAm measurements. A previous study of incident type 2 diabetes reported methylation at 2 out of their 5 discovered CpG sites were correlated across blood and liver.^18^ Our sample size in White adults was smaller and we may have been underpowered to discover potentially novel loci in this population. Although we sought to capture changes in DNAm that lead to hyperglycemia by performing a prospective analysis of individuals without diabetes, pathogenesis of type 2 diabetes and hyperglycemia may precede diagnosis by several years.^77^ We did not perform any *in silico* or functional follow up to confirm if the CpG site associations we identified were truly causes or consequences of type 2 diabetes. A longitudinal study with a longer follow-up period could be advantageous in detecting whether DNAm changes in later life type 2 diabetes appear early in life since early childhood is a sensitive period for DNAm changes to occur in response to adversity^78^. We were unable to tease apart if differences observed between groups were reflective of differences in geography or race since Black adults were solely from Jackson, Mississippi, whereas White adults were primarily from Forsyth, North Carolina. Additionally, we only had DNAm measurement from a single time point and were unable to consider it as a time-varying exposure in our time-to-event analysis. We did not replicate our findings in an external population of Black and White adults to assess generalizability of our results. While we examined explained variance of type 2 diabetes using DNAm levels from significant CpG sites in addition to traditional type 2 diabetes risk factors, we were not able to evaluate a predictive model due to lack of an external dataset to validate such a model.

In conclusion, our study exploring diverse US populations revealed novel DNAm-type 2 diabetes associations including CpG sites at genes such as *GPX6* and *JPH3*, and a differentially methylated gene, *TP63* previously linked to diabetes in experimental studies. Further gene-specific DNAm studies and environmental factors upstream to identified CpG sites can help elucidate the role of epigenetics in the pathogenesis of type 2 diabetes.

## SUPPLEMENTAL DATA

Supplemental Data includes additional methodological details (Supplementary Methods) and figures (Supplementary Figures). Supplementary Tables are available in a separate excel document.

## Data Availability

ARIC has a substantial amount of data in repositories such as BioLINCC and dbGaP. Not all individual-level ARIC data are publicly available because of consent restrictions. All summary-level data produced in the present work are contained in the manuscript. Other data may be available upon reasonable request to the authors.

## ACKNOWLEDGEMENTS

The analyses performed in this manuscript was carried out using computational resources of the Joint High Performance Computing Exchange (JHPCE) maintained by the Department of Biostatistics at the Johns Hopkins University. The authors thank the staff and participants of the ARIC study for their important contributions.

## AUTHOR CONTRIBUTIONS

Conceptualization: SV, DR

Data curation: SV

Formal analysis: SV

Funding acquisition: DR

Investigation: SV

Methodology: SV, DR, JSP

Project administration: ES, DR

Supervision: JSP, ES, DR

Validation: SV

Visualization: SV

Writing – original draft: SV

Writing – review & editing: SV, JSP, EB, MF, ES, DR

