## Supplementary Text and Figures for "Epigenome-wide association study of incident type 2 diabetes in Black and White participants from the Atherosclerosis Risk in Communities Study"

### SUPPLEMENTARY METHODS

*Measurement of DNAm Levels*

**Bisulfite conversion.** DNA was extracted from peripheral blood leukocyte samples using Gentra Puregene Blood Kit (Qiagen) and bisulfite conversion was done using EZ-96 DNA Methylation Kit (Deep Well Format). Further details are available elsewhere.[1]

**Measurement and quality control.** DNAm levels at individual CpG sites were measured using the Illumina Infinium HumanMethylation450 BeadChip array also known as the HM450K array. This array assays >480,000 CpG sites covering approximately 99% of RefSeq genes and 96% of CpG islands among other regions with multiple probes per region.[2] Degree of methylation was determined using Illumina GenomeStudio 2011.1 Methylation module 1.9.0 software. Background correction was performed by subtracting channel-specific background, calculated from negative controls in a given channel, from probe intensities in the corresponding channel. DNAm levels at each CpG site, represented as beta ($\beta$) values with range 0-1 (0 is non-methylated and 1 is completely methylated), was estimated as the ratio of intensity of the methylated probe to the intensity of the methylated probe + unmethylated probe. Sample-level and CpG site-level quality control (QC) steps were undertaken. Sample-level QC included filtering out samples with 1) failed bisulfite conversion, 2) gender discordance determined by matching Y chromosome probes with Exome Chip or MetaboChip data, 3) SNP mismatch between SNPs available on the HM450K array and existing genotype data from other arrays/chips, 4) poor clustering by sex in MDS plots, 5) low probe intensity, and 6) ≥1% missing data. A total of 2,796 Black and 1,139 White participants were retained after sample-level QC. In Black adults, additional CpG site-level QC were applied including filtering out CpG sites with bead count <3 in ≥5% of samples and CpG sites with ≥1% missing data. For our analyses, we examined CpG sites on autosomes only, which resulted in 470,161 and 469,973 CpG sites for Black adults and White adults respectively. All genomic coordinates are given in NCBI Build GRCh37/UCSC hg19.

**Normalization.** The HM450K array consists of two probe types with differences in design that can lead to variability in β-values measured by them. Following background correction and QC, Beta Mixture Quantile dilation (BMIQ) normalization was performed to ensure β-values from Infinium II probes have a distribution similar to Infinium I probes. BMIQ normalization reduces technical variation, bias of Infinium II probe values due to β-values not detecting values closer to the ends of 0-1 range, and enrichment bias of Infinium I probe.[3]

*Measurement of Cell Type Proportions*

Since we have blood-based DNAm measurements and blood consists of multiple cell types with heterogenous DNAm profiles, we need to adjust for cell composition variability.[4] A white blood cell (WBC) differential was measured in 175 Black participants during baseline visit from blood specimens drawn and stored at 4°C in hospital-based, independent laboratories within 24 hours after venipuncture.[5, 6] WBC proportions of neutrophils, lymphocytes, monocytes, eosinophils and basophils were imputed in remaining Black participants using the measured subset as reference and the imputation algorithm proposed by Houseman et. al.[7, 8] Basophils had low imputation quality (leave-one-out approach was used to evaluate the cell type imputation accuracy) and were excluded due to low percentage of overall differential (0-2%). Cell type proportions in White participants were estimated using the `estimateCellCounts` function in `minfi` R package[9], which provides composition of specific white blood cell types only (B, $CD4^+$ T, $CD8^+$ T, eosinophils, granulocytes, monocytes, neutrophils, and natural killer cells) based on its in-built HM450K reference dataset only available for Europeans. The ARIC data on White adults had cell type proportions estimated for B, $CD4^+$ T, $CD8^+$ T, granulocytes, monocytes, and NK. All our analyses were race-stratified and then combined in meta-analysis.

*Statistical Analyses*

**Time-to-event analysis using secondary models satisfying proportional hazards assumption.** Besides the primary model (Model 1) with covariate adjustments for age, sex, smoking status, education level, and the first 10 genetic PCs, we additionally fit secondary models to assess if significant CpG sites from Model 1 were associated with incident type 2 diabetes independent of BMI and fasting glucose. BMI is a major risk factor for type 2 diabetes[10] and a potential confounder for the association of DNAm levels at CpG sites with type 2 diabetes. It is also possible that some CpG sites influence incident type 2 diabetes risk through BMI-related pathways. So, our Model 2 included all covariates from Model 1 along with BMI as a continuous covariate and BMI x time interaction term in both race groups to model potential BMI-time dependence and ensure the proportional hazards assumption was met. To examine if the significant CpG sites from Model 1 were related to conversion to T2D or just marking subtle differences in hyperglycemia that are already present at baseline, our Model 3 included all covariates from Model 1 along with time-varying fasting glucose effects that differed across periods of time since time origin. Unlike BMI, adjusting fasting glucose as a covariate with effect changing linearly with time did not satisfy proportional hazards assumption. Allowing for differing fasting glucose effects by two evenly split time periods in Black adults and eleven evenly split time periods in White adults satisfied the proportional hazards assumption. Thereafter, we meta-analyzed race-stratified results for each model to examine if the significant CpG sites remained statistically significant after BMI or fasting glucose adjustment and if there was any attenuation in log hazard ratios. For this, we calculated percent change in effect size as $100 \times (\log HR_{\text{no-BMI-adj}} - \log HR_{\text{BMI-adj}}) / \log HR_{\text{no-BMI-adj}}$ (similarly for fasting glucose), where a positive value is indicative of attenuation in effect size due to adjustment. A 0-3% change in effect size was considered minimal change.[11]

**Sensitivity analysis for genetic relatedness.** We did not remove related individuals in any of our analyses because on average heritability estimates across genome-wide DNAm sites are low[12]. To assess if removal of relatives qualitatively influenced our results, we first identified first-degree relatives as pairs of individuals with kinship coefficient of $\geq 2^{-5/2}$ using KING-robust estimator.[13] We then used `pcairPartition`[14] from R package GENESIS[15] to retain a set of unrelated samples that were most representative of ancestry where no pair of samples in the unrelated subset were first-degree relatives. The unrelated subset contained 1,979 Black and 1,019 White adults. We fit our primary model on this unrelated subset and compared findings with those from all individuals.

**Sensitivity analysis for outcome definition.** We sought to understand if findings from the primary model were consistent across different definitions of type 2 diabetes. For this sensitivity analysis, we defined incident type 2 diabetes cases based on self-reported medication use only (a more stringent definition of type 2 diabetes) because cases of doctor diagnosis may be over-reported.

**Sensitivity analysis for proportional hazards assumption.** We compared signals from the BMI-adjusted and the fasting glucose-adjusted secondary models with and without the proportional hazards assumption being met. Specifically, for the BMI adjusted model, we examined if the same signals were identified from our Model 2 that included both BMI and BMI x time interaction terms (which satisfied the proportional hazards assumption) and from a simpler model including BMI as a continuous variable (which failed the proportional hazards assumption). For fasting glucose, we similarly compared findings from our Model 3 against those from a simpler model that included fasting glucose as a continuous variable.

**Differentially methylated regions.** We conducted differentially methylated region (DMR) analysis across two broadly defined categories.

*Gene-region specific DMR analysis:* Gene-region specific DMRs were identified by analyzing grouped CpG sites within each gene region including 1500bp ahead of transcription start site (TSS1500), 200 bp ahead of transcription start site (TSS200), 1$^{st}$ exon of gene (Exon1), gene body regions post 1$^{st}$ exon (Genebody), and 3' untranslated region (3'UTR). Gene-regions of TSS1500, TSS200, Exon1, Genebody, and 3'UTR were identified using `fullannotInd` dataset based on Illumina methylation annotation available through the R package `IMA`.[16] CpG sites annotated to each gene-region were grouped together and the region was defined based on the boundaries set by chromosomal positions of the CpG sites. Most gene-regions spanned ≤1Mb with a few extreme outliers falling outside this range. We restricted the gene-region specific DMR analysis to regions spanning ≤1Mb to reduce the computational burden of the auto-correlation function (ACF) calculation (details provided below). We examined a total of 20,403 TSS1500, 17,728 TSS200, 15,528 Exon1, 19,024 Genebody, and 14,072 UTR3 gene-regions in the region-specific DMR analysis. The gene-region size distribution is depicted in **Supplementary Figure 1**. We implemented DMR analysis using `comb-p`[17] (a summary statistics-based method) on the individual CpG site p-values from our primary model analysis. First, the ACF was calculated for adjacent p-values extending to the length of the longest region with a step-size of 50. Second, the Stouffer-Liptak-Kechris correction was applied to obtain p-value for each region. Finally, the p-value for each region was adjusted for multiple-testing correction by calculating the Šidák-corrected p-value, where the total number of tests in a region is the total number of bases divided by the length of a given region.

*Gene-region agnostic DMR analysis:* Gene-region agnostic DMRs were regions of CpG site associations in close proximity (within 1 to 500 bases) identified regardless of well-defined gene or gene-region boundaries. To perform gene-agnostic DMR analysis, an initial ACF was calculated for adjacent p-values within 1 to 500 bases with a step-size of 50. Second, the estimated ACF was used to adjust the p-values using the Stouffer-Liptak-Kechris correction. Third, the corrected p-values were used to identify peaks or regions of low p-values using a seed p-value of 0.01 the region was further extended to include any additional CpG sites within 500 Kb. Once the regions with p-value peaks were identified, a second ACF was calculated accounting for the length of the longest peak and the Stouffer-Liptak-Kechris correction was applied again to obtain p-value for each region. Following the Stouffer-Liptak-Kechris correction, a multiple testing corrected Šidák p-value was obtained for each region.[18, 19]

#### SUPPLEMENTARY FIGURES

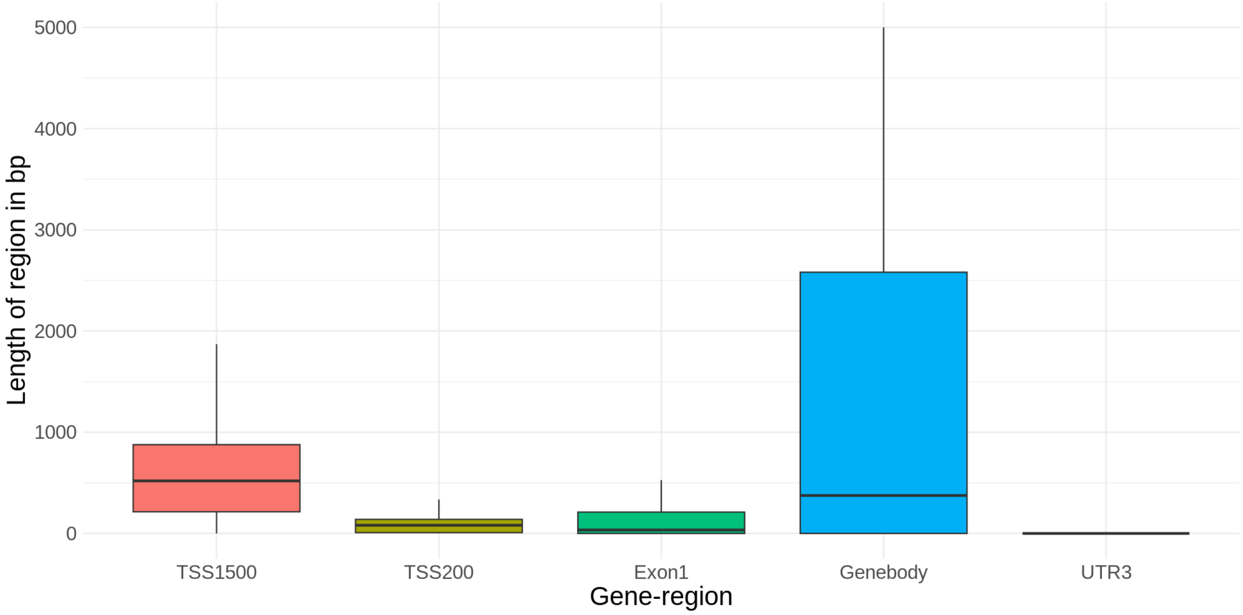


**Supplementary Figure 1:** Boxplot of predefined gene-region length used in gene-region specific DMR analysis after filtering for regions with length ≤1Mb.

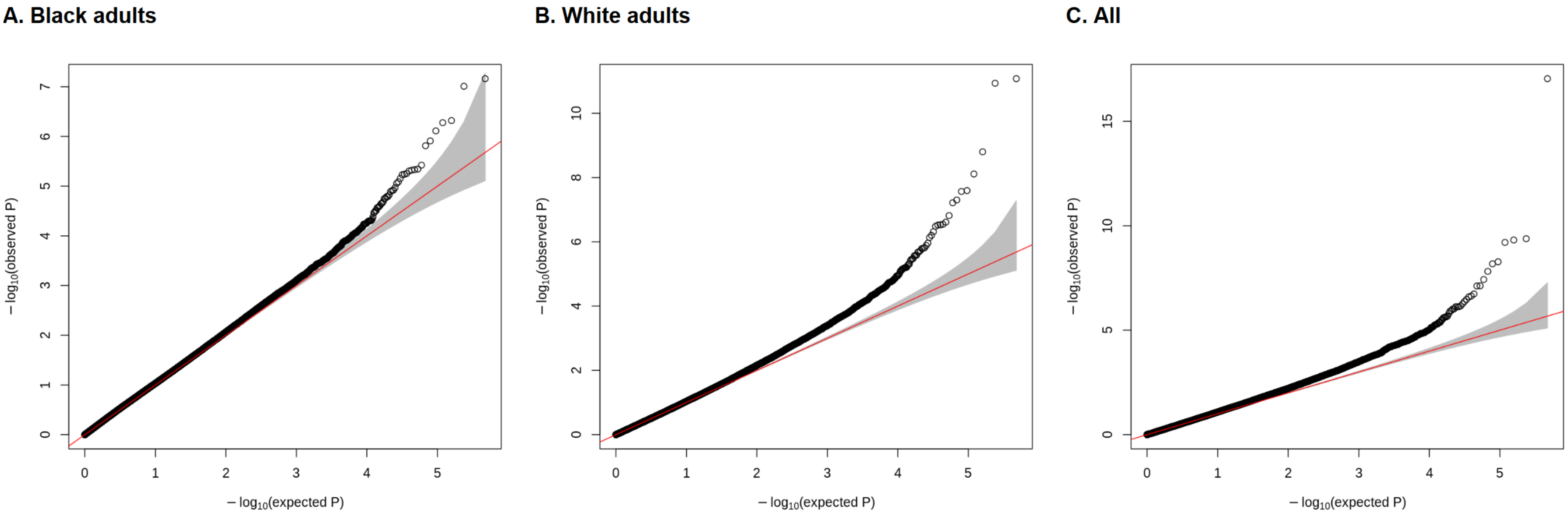


**Supplementary Figure 2:** QQ plots of epigenome-wide CpG site analysis of incident type 2 diabetes using Cox proportional hazards model adjusting for age, sex, smoking status, education level, and the first 10 genetic principal components. Technical covariates (chip ID, chip row, study center, visit, project) and cell type proportions were adjusted too. The gray shaded region represents a conservative 95% confidence interval for the expected distribution of p-values.

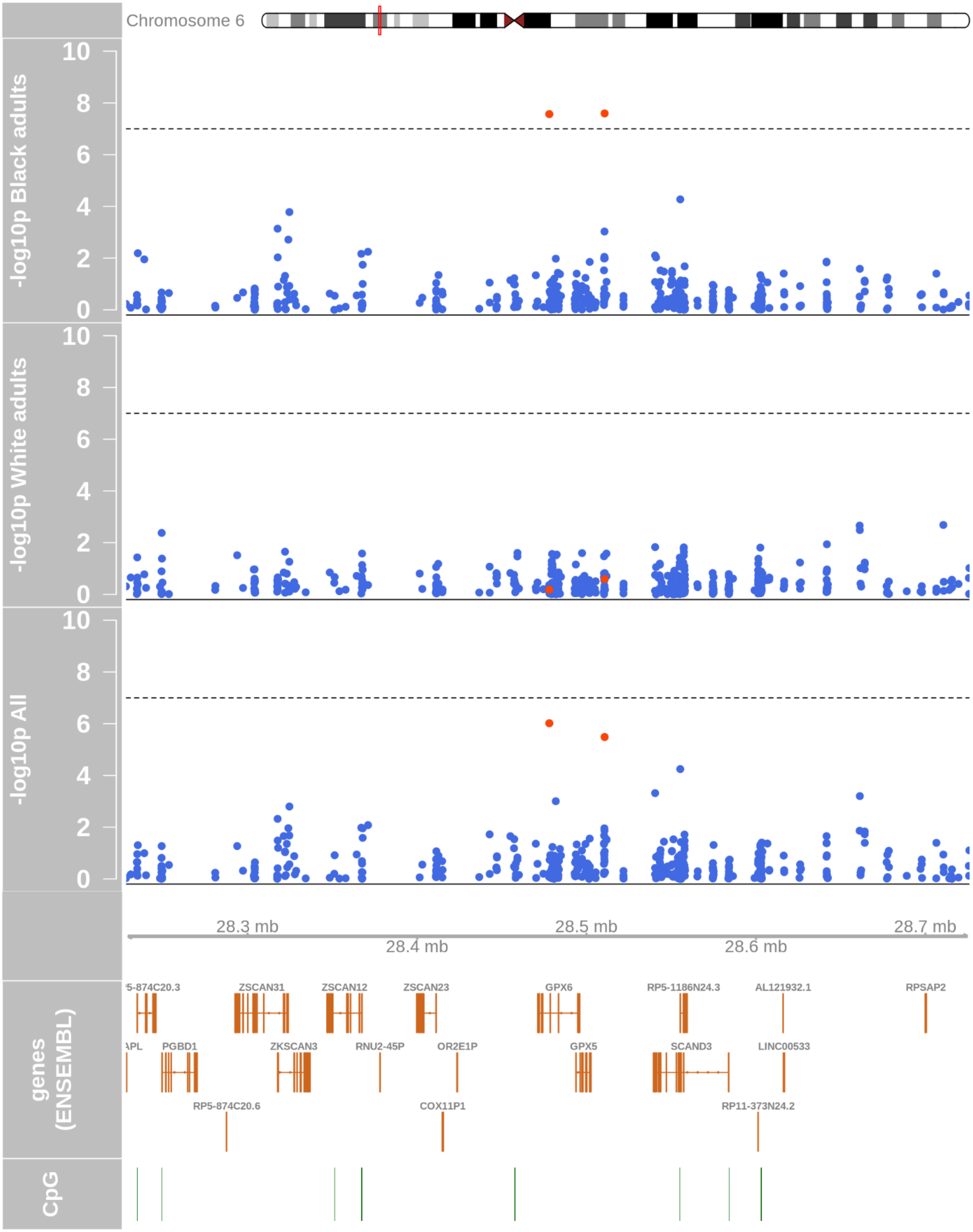


**Supplementary Figure 3:** Regional association plot of CpG sites at chr6p22 near *GPX6* in an epigenome-wide incident type 2 diabetes analysis across race groups. A Cox proportional hazards model adjusting for age, sex, smoking status, education level, and the first 10 genetic principal components was fit. Technical covariates (chip ID, chip row, study center, visit, project) and cell type proportions were adjusted too. Negative log-transformed p-values of association are plotted against chromosome coordinates, genes and CpG islands. Black dashed lines correspond to Bonferroni-corrected epigenome-wide significance threshold of $10^{-7}$.

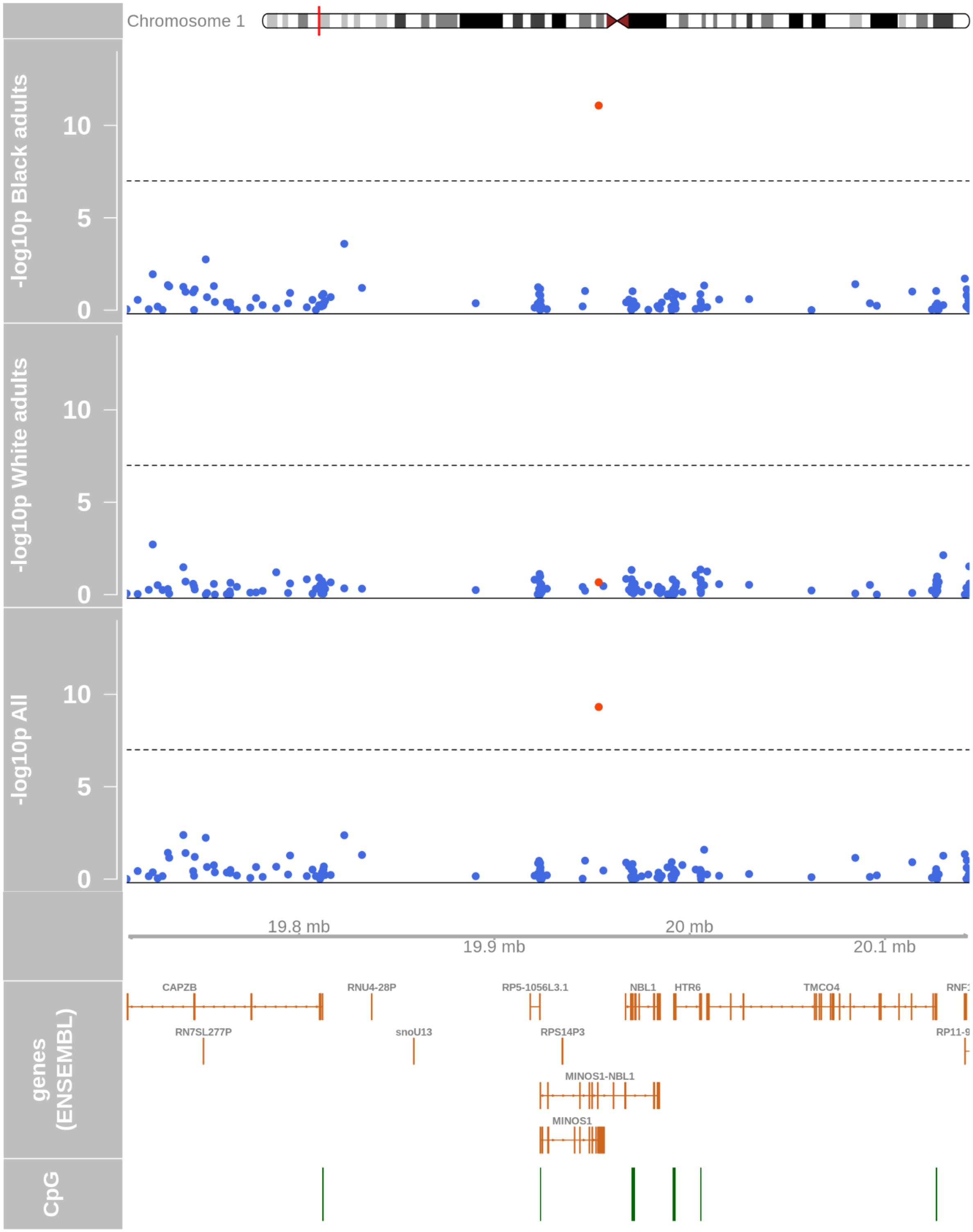


**Supplementary Figure 4**: Regional association plot of CpG sites at chr1q21 near *C1orf151* (or *MINOS1*) in an epigenome-wide incident type 2 diabetes analysis across race groups. A Cox proportional hazards model adjusting for age, sex, smoking status, education level, and the first 10 genetic principal components was fit. Technical covariates (chip ID, chip row, study center, visit, project) and cell type proportions were adjusted too. Negative log-transformed p-values of association are plotted against chromosome coordinates, genes and CpG islands. Black dashed lines correspond to Bonferroni-corrected epigenome-wide significance threshold of $10^{-7}$.

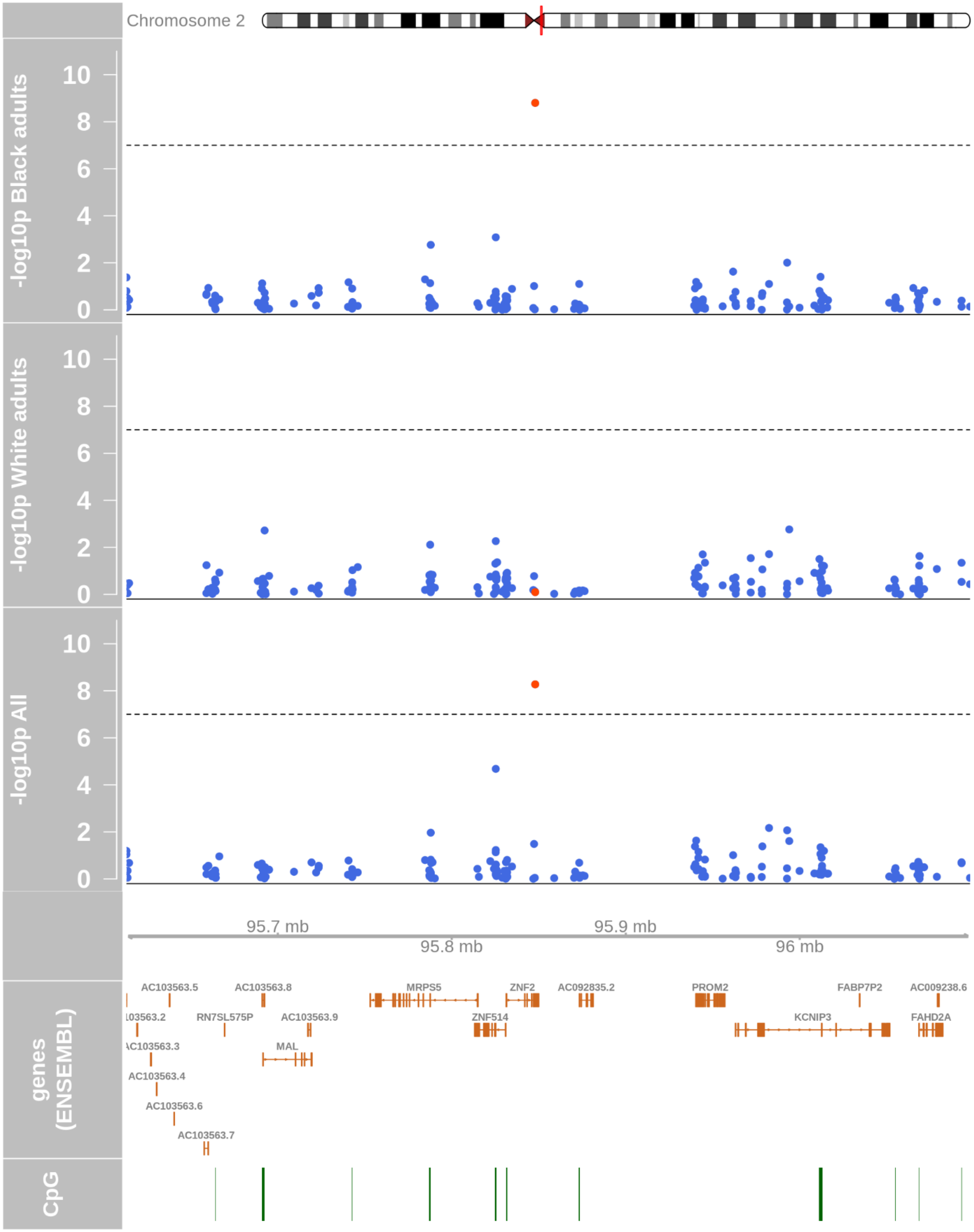


**Supplementary Figure 5**: Regional association plot of CpG sites at chr2q11 near *ZNF2* in an epigenome-wide incident type 2 diabetes analysis across race groups. A Cox proportional hazards model adjusting for age, sex, smoking status, education level, and the first 10 genetic principal components was fit. Technical covariates (chip ID, chip row, study center, visit, project) and cell type proportions were adjusted too. Negative log-transformed p-values of association are plotted against chromosome coordinates, genes and CpG islands. Black dashed lines correspond to Bonferroni-corrected epigenome-wide significance threshold of $10^{-7}$.

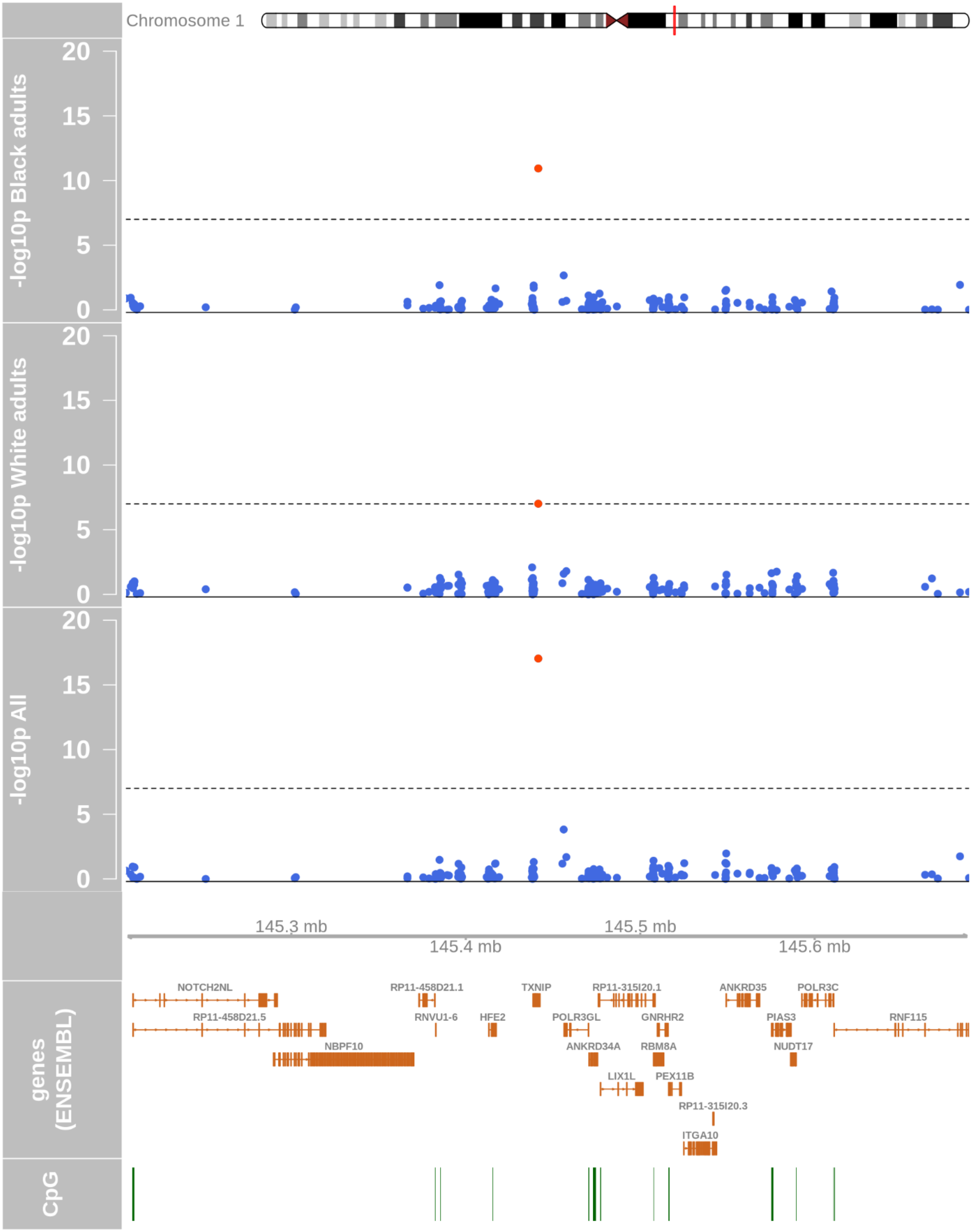


**Supplementary Figure 6**: Regional association plot of CpG sites at chr1q21 near *TXNIP* in an epigenome-wide incident type 2 analysis across race groups. A Cox proportional hazards model adjusting for age, sex, smoking status, education level, and the first 10 genetic principal components was fit. Technical covariates (chip ID, chip row, study center, visit, project) and cell type proportions were adjusted too. Negative log-transformed p-values of association are plotted against chromosome coordinates, genes and CpG islands. Black dashed lines correspond to Bonferroni-corrected epigenome-wide significance threshold of $10^{-7}$.

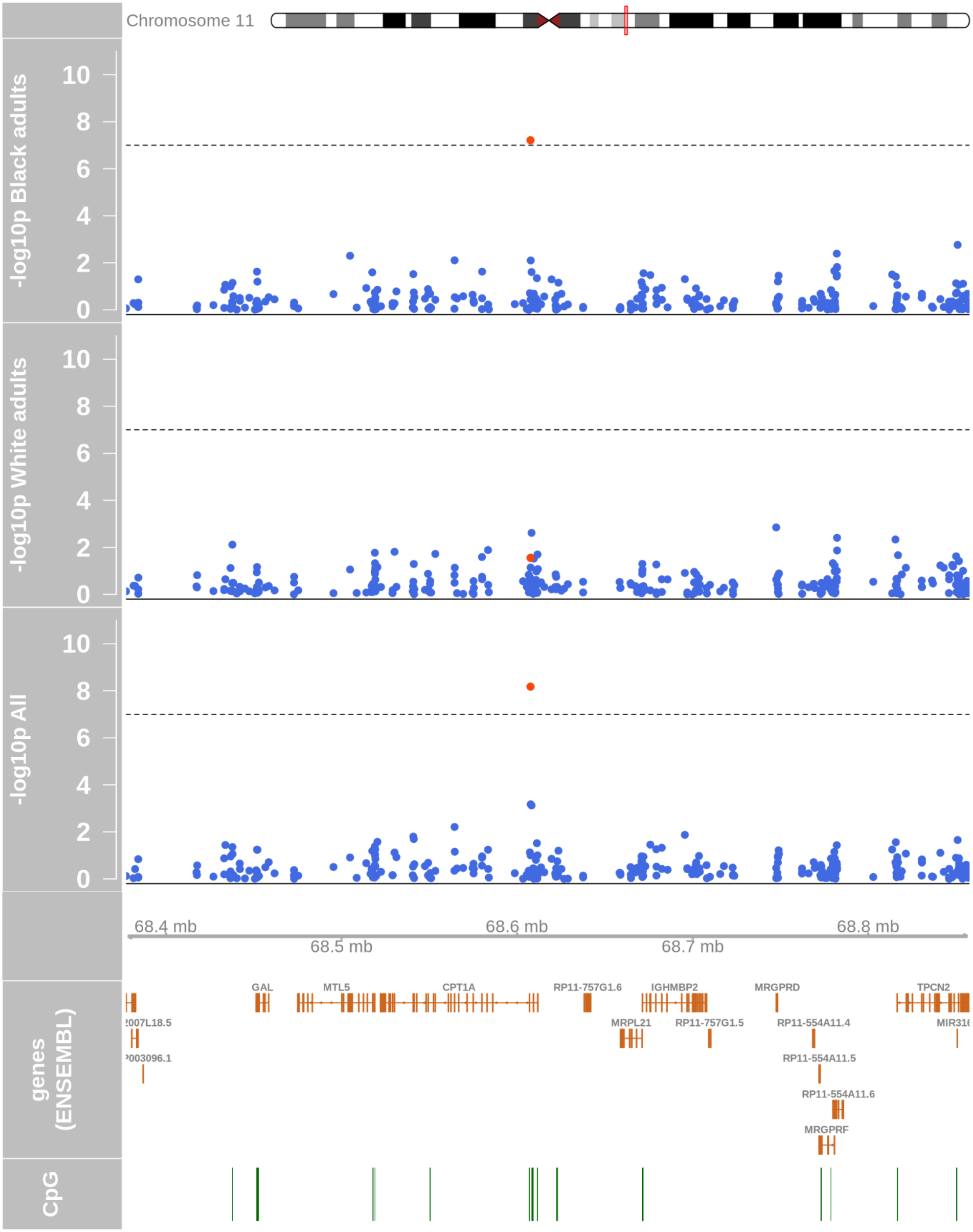


**Supplementary Figure 7**: Regional association plot of CpG sites at chr11q13 near *CPT1A* in an epigenome-wide incident type 2 analysis across race groups. A Cox proportional hazards model adjusting for age, sex, smoking status, education level, and the first 10 genetic principal components was fit. Technical covariates (chip ID, chip row, study center, visit, project) and cell type proportions were adjusted too. Negative log-transformed p-values of association are plotted against chromosome coordinates, genes and CpG islands. Black dashed lines correspond to Bonferroni-corrected epigenome-wide significance threshold of $10^{-7}$.

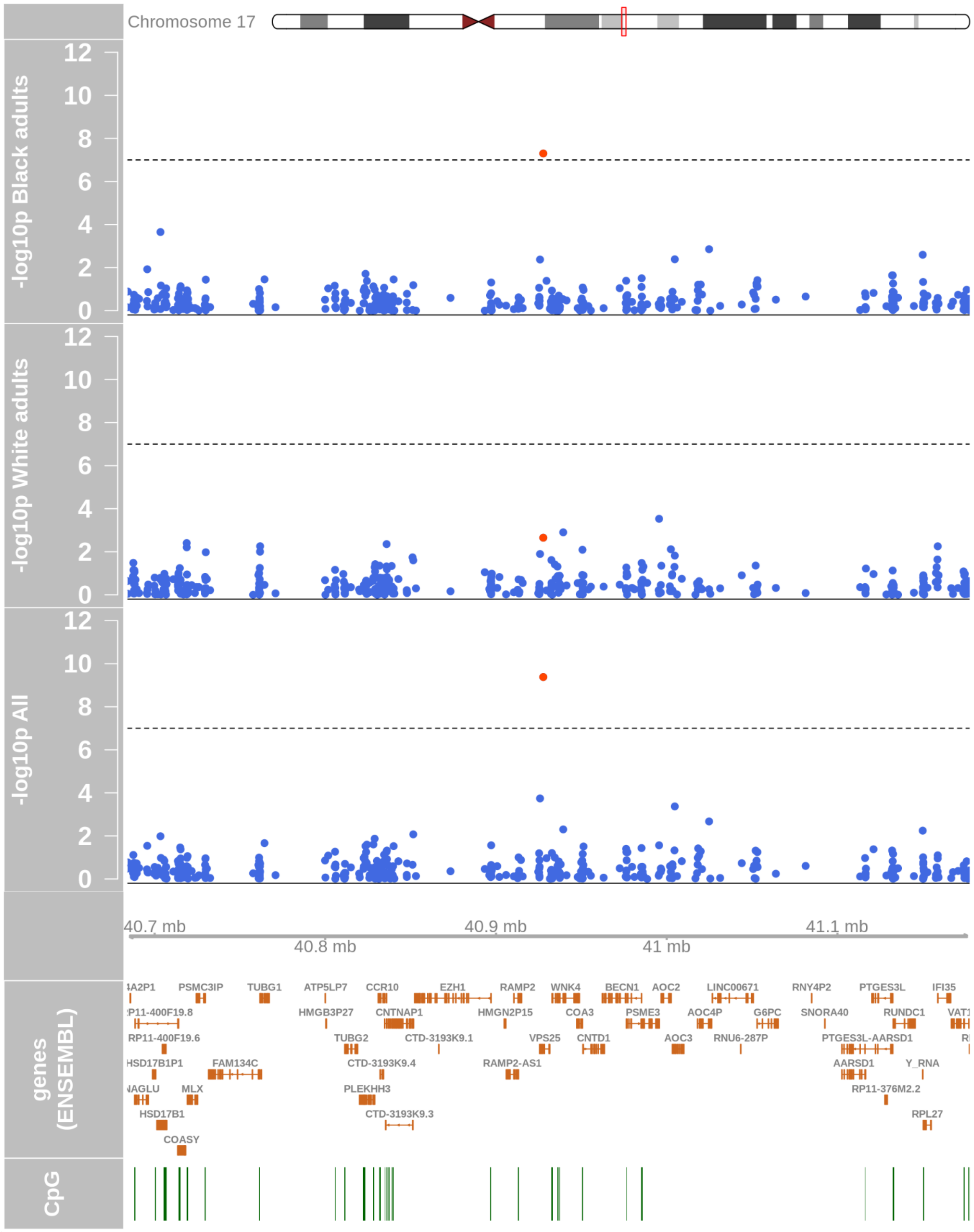


**Supplementary Figure 8**: Regional association plot of CpG sites at chr17q21 near *VPS25* in an epigenome-wide incident type 2 analysis across race groups. A Cox proportional hazards model adjusting for age, sex, smoking status, education level, and the first 10 genetic principal components was fit. Technical covariates (chip ID, chip row, study center, visit, project) and cell type proportions were adjusted too. Negative log-transformed p-values of association are plotted against chromosome coordinates, genes and CpG islands. Black dashed lines correspond to Bonferroni-corrected epigenome-wide significance threshold of $10^{-7}$.

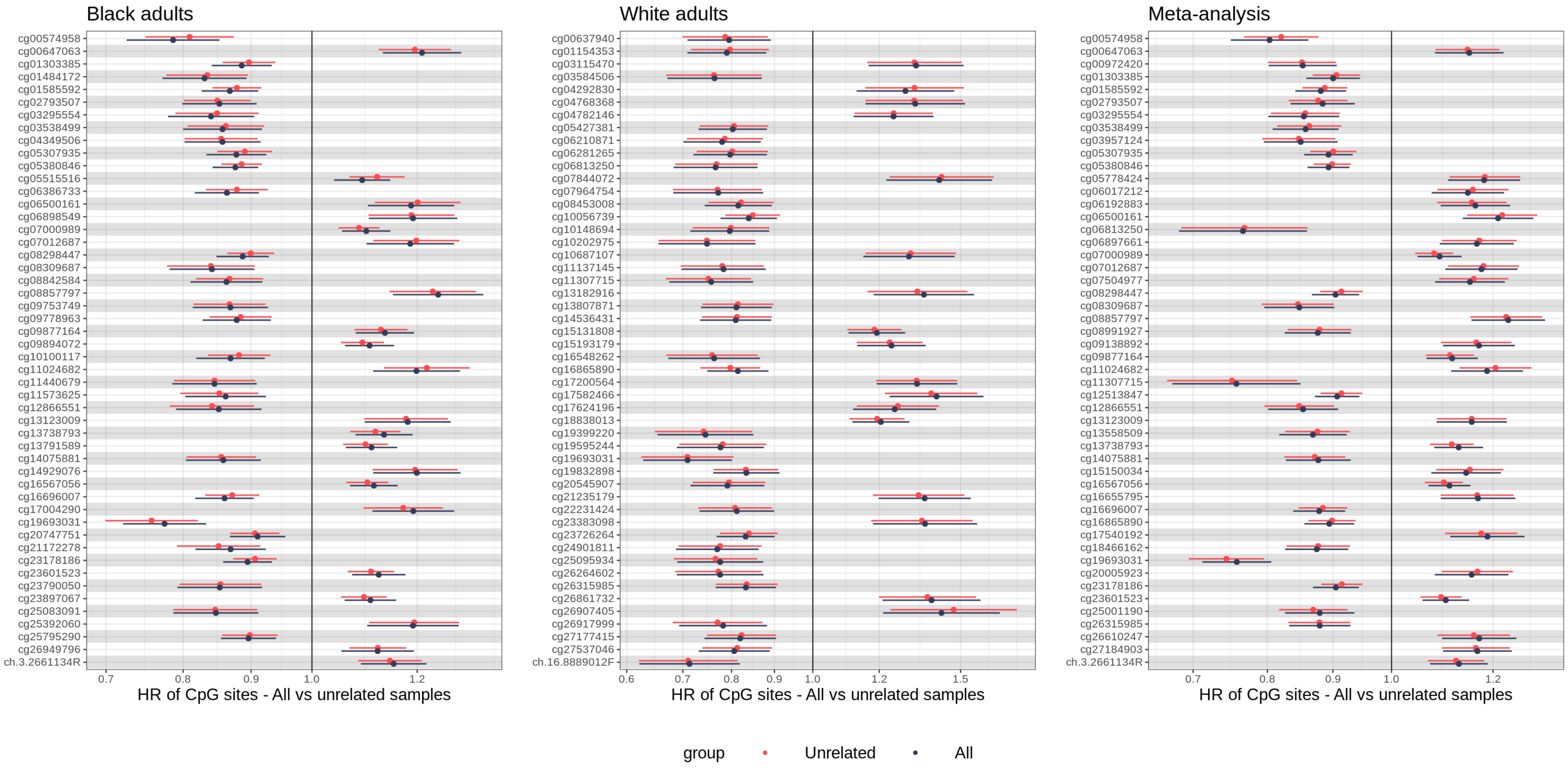


**Supplementary Figure 9**: Forest plot of effect sizes and their 95% confidence intervals for the top 50 CpG site associations across race groups to assess their sensitivity to related individuals in the sample. The effect estimates were obtained from Cox proportional hazards model adjusting for age, sex, smoking status, education level, and the first 10 genetic principal components. Technical covariates (chip ID, chip row, study center, visit, project) and cell type proportions were adjusted too. The 'All' group included all participants (2,091 Black and 1,029 White adults) while the 'Unrelated' group included participants after removing first-degree relatives (1,979 Black and 1,019 White adults).

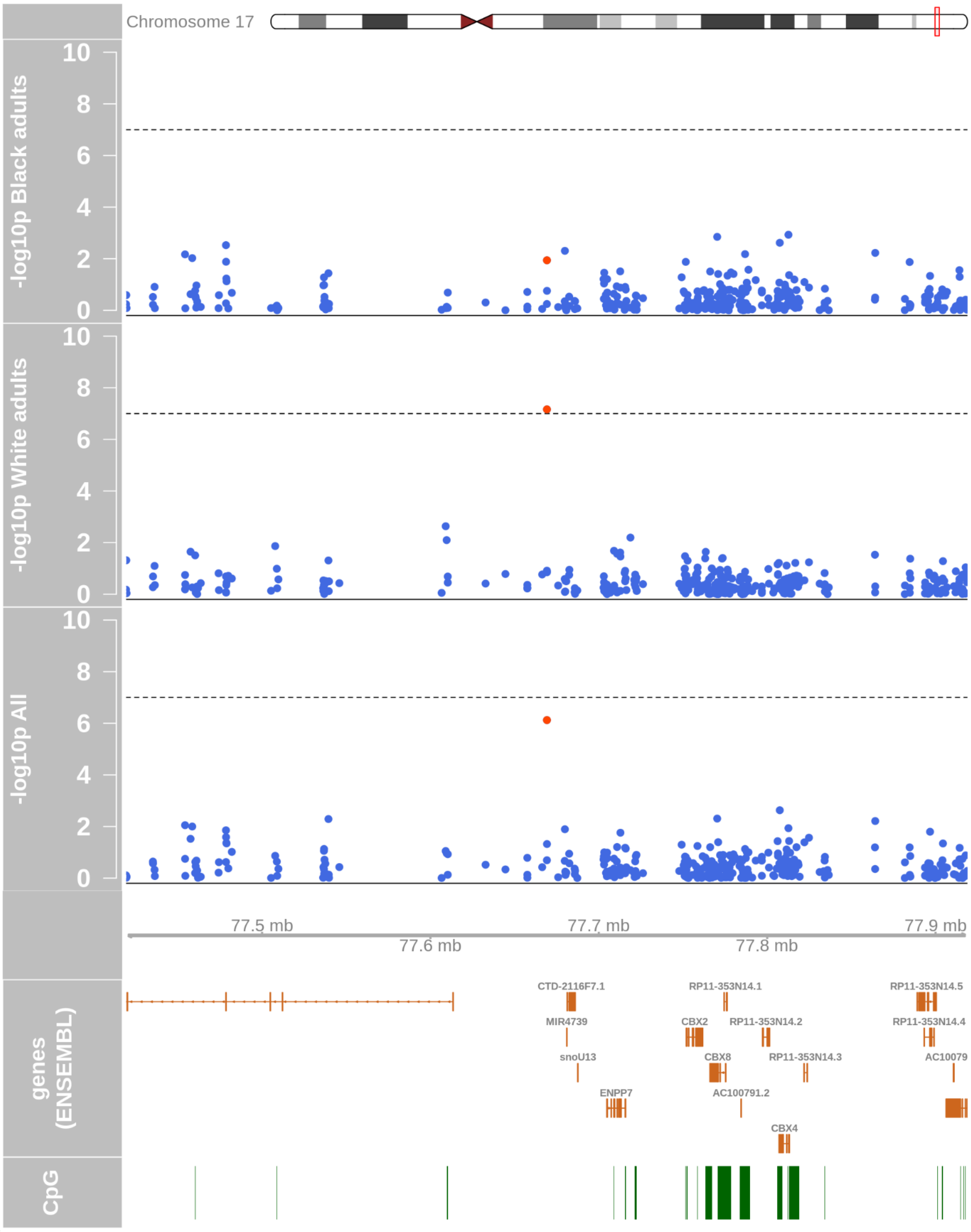


**Supplementary Figure 10**: Regional association plot of CpG sites at chr17q25 in an epigenome-wide incident type 2 analysis across race groups. A Cox proportional hazards model adjusting for age, sex, smoking status, education level, and the first 10 genetic principal components was fit. Technical covariates (chip ID, chip row, study center, visit, project) and cell type proportions were adjusted too. Negative log-transformed p-values of association are plotted against chromosome coordinates, genes and CpG islands. Black dashed lines correspond to Bonferroni-corrected epigenome-wide significance threshold of $10^{-7}$.

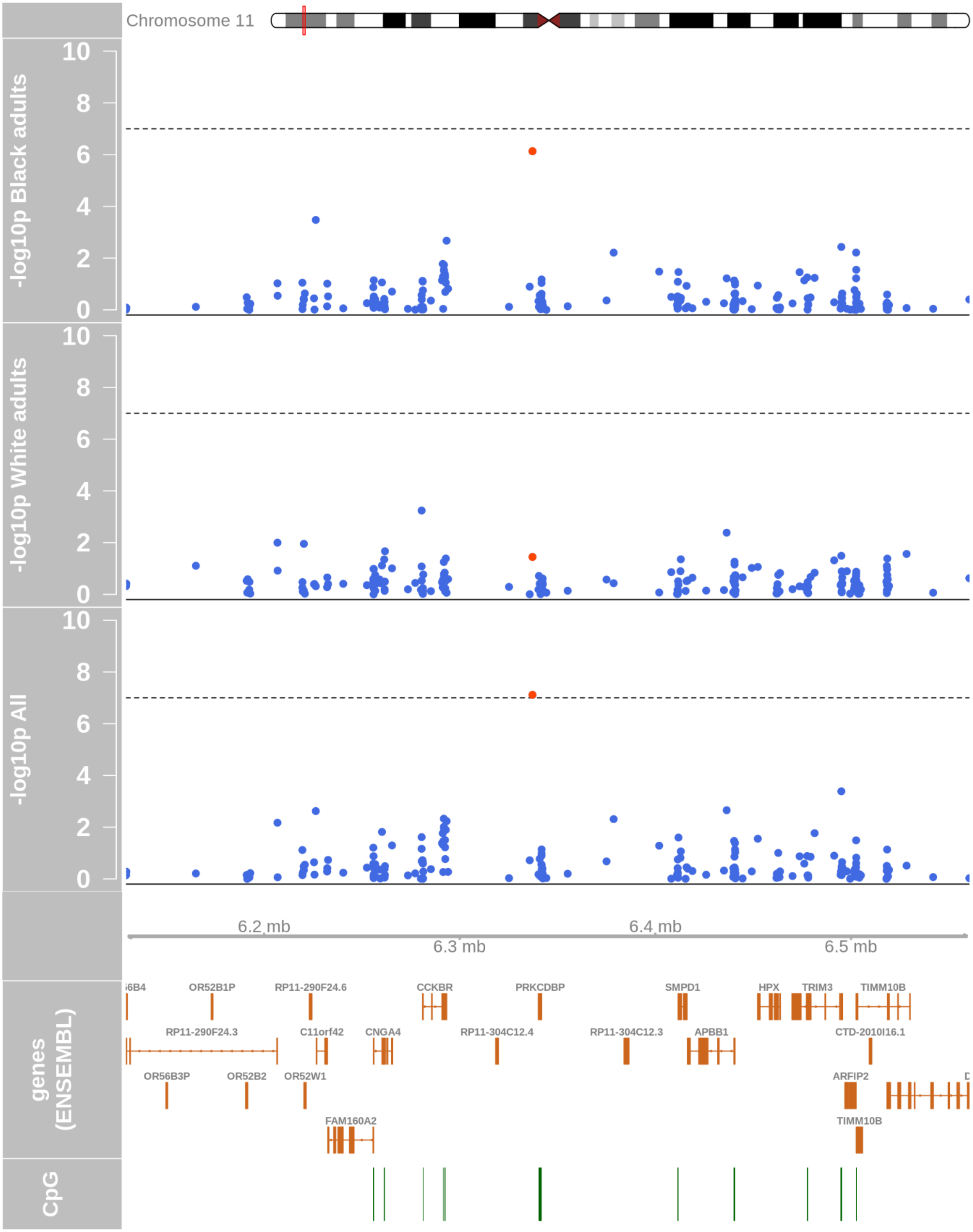


**Supplementary Figure 11**: Regional association plot of CpG sites at chr11p15 in an epigenome-wide incident type 2 analysis across race groups. A Cox proportional hazards model adjusting for age, sex, smoking status, education level, and the first 10 genetic principal components was fit. Technical covariates (chip ID, chip row, study center, visit, project) and cell type proportions were adjusted too. Negative log-transformed p-values of association are plotted against chromosome coordinates, genes and CpG islands. Black dashed lines correspond to Bonferroni-corrected epigenome-wide significance threshold of $10^{-7}$.

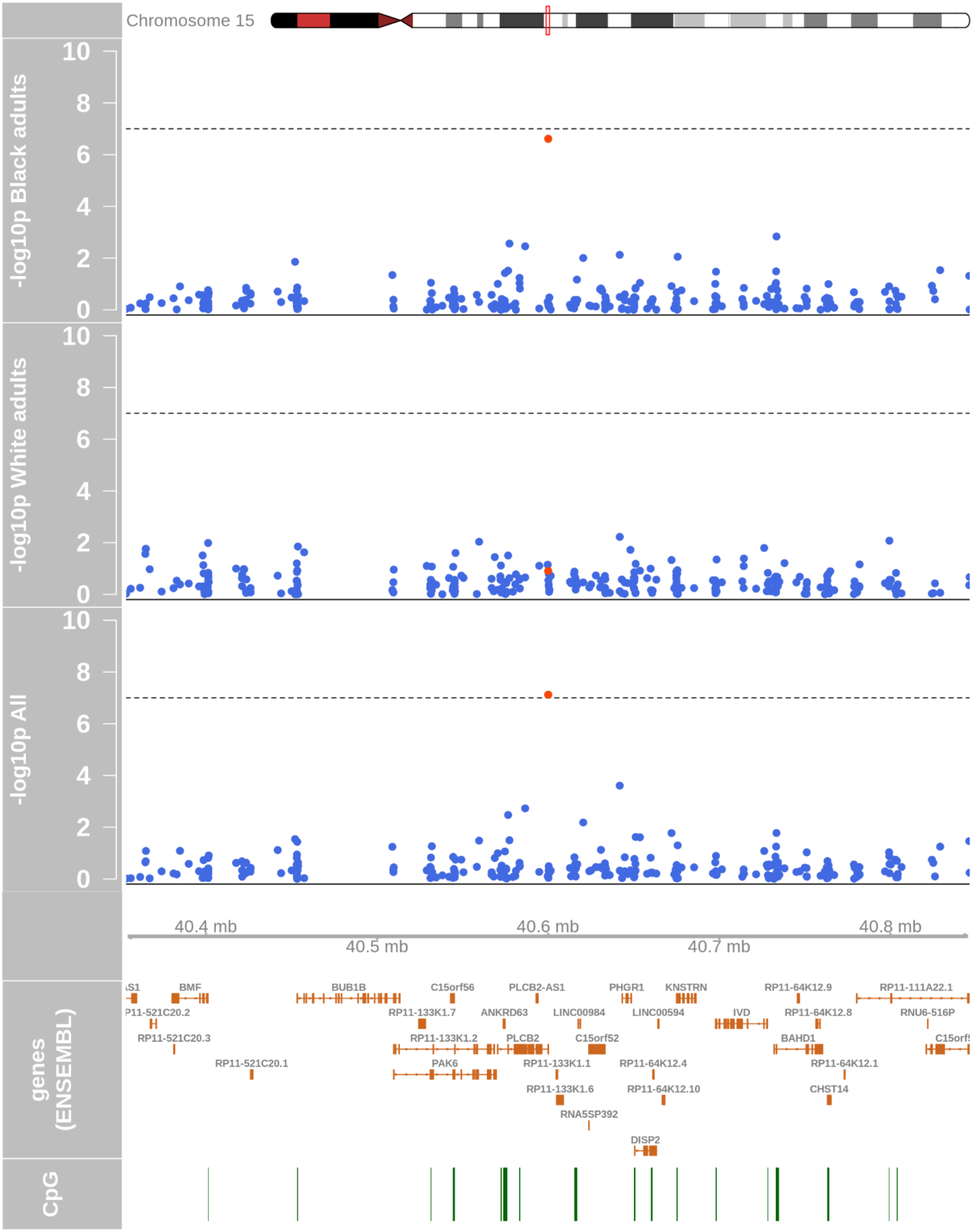


**Supplementary Figure 12**: Regional association plot of CpG sites at chr15q15 near *PLCB2* in an epigenome-wide incident T2D analysis across race groups. A Cox proportional hazards model adjusting for age, sex, smoking status, education level, and the first 10 genetic principal components was fit. Technical covariates (chip ID, chip row, study center, visit, project) and cell type proportions were adjusted too. Negative log-transformed p-values of association are plotted against chromosome coordinates, genes and CpG islands. Black dashed lines correspond to Bonferroni-corrected epigenome-wide significance threshold of $10^{-7}$.

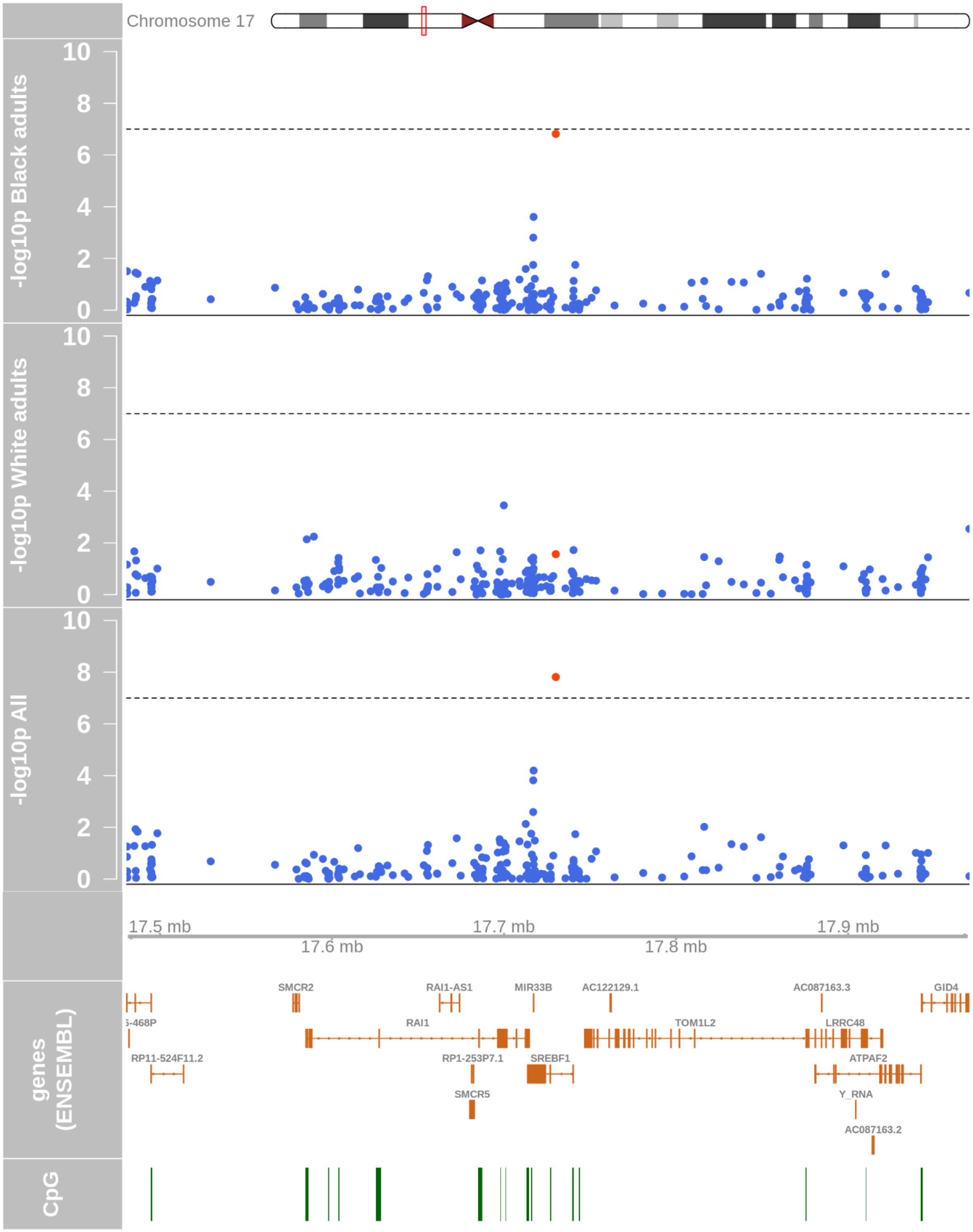


**Supplementary Figure 13**: Regional association plot of CpG sites at chr17p11 near *SREBF1* in an epigenome-wide incident type 2 diabetes analysis across race group. A Cox proportional hazards model adjusting for age, sex, smoking status, education level, and the first 10 genetic principal components was fit. Technical covariates (chip ID, chip row, study center, visit, project) and cell type proportions were adjusted too. Negative log-transformed p-values of association are plotted against chromosome coordinates, genes and CpG islands. Black dashed lines correspond to Bonferroni-corrected epigenome-wide significance threshold of $10^{-7}$.

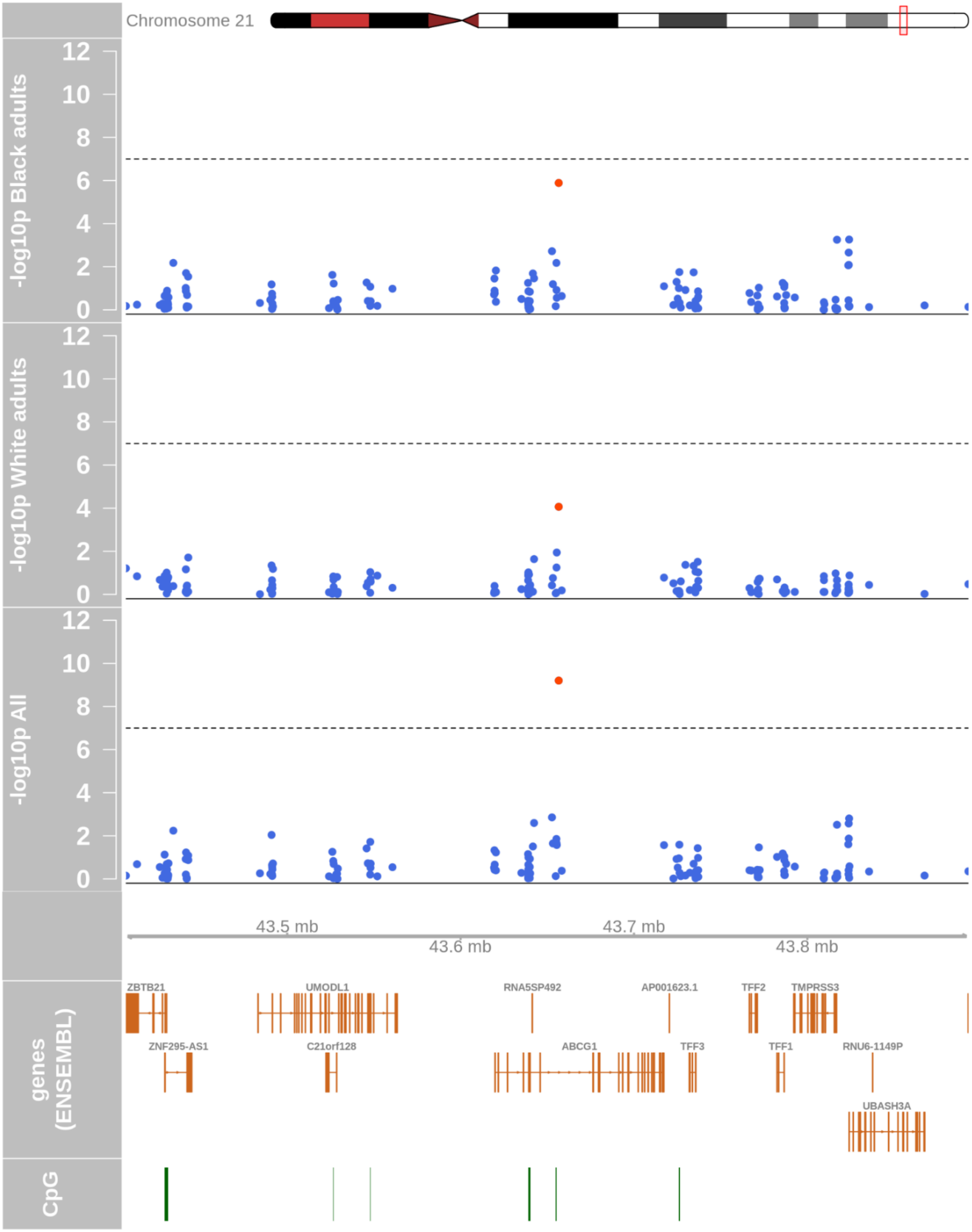


**Supplementary Figure 14**: Regional association plot of CpG sites at chr21q22 near *ABCG1* in an epigenome-wide incident type 2 diabetes analysis across race groups. A Cox proportional hazards model adjusting for age, sex, smoking status, education level, and the first 10 genetic principal components was fit. Technical covariates (chip ID, chip row, study center, visit, project) and cell type proportions were adjusted too. Negative log-transformed p-values of association are plotted against chromosome coordinates, genes and CpG islands. Black dashed lines correspond to Bonferroni-corrected epigenome-wide significance threshold of $10^{-7}$.

**A. Black adults** **B. White adults** **C. All**

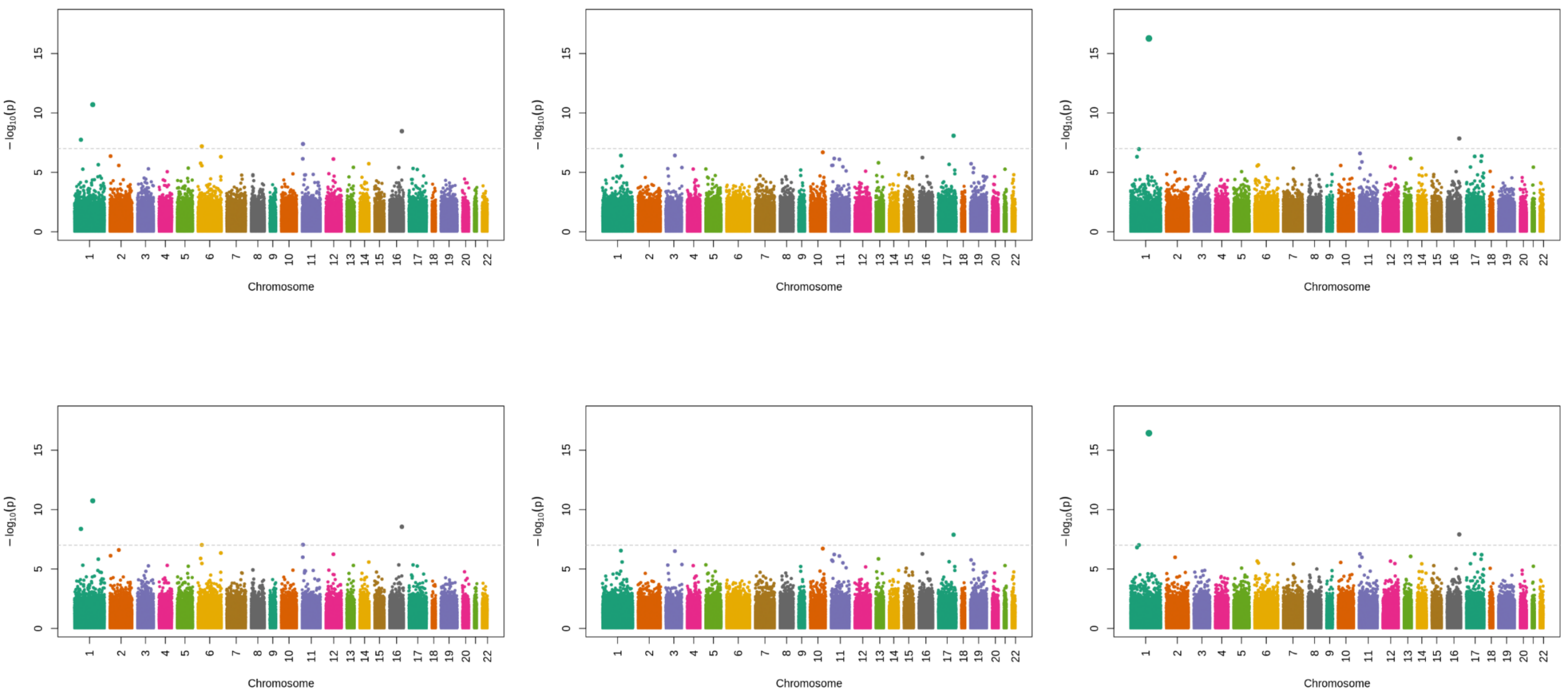


**Supplementary Figure 15**: Manhattan plots for BMI-adjusted Cox proportional hazards models showing differences in DNAm associations with and without violation of proportional hazards (PH) assumption. For each race group, the top Manhattan plot shows the CpG site p-values from the Cox proportional hazards model adjusted for age, sex, smoking status, education level, BMI along with BMI x time interaction, and the first 10 genetic principal components (this model satisfies the PH assumption). The bottom Manhattan plot shows the CpG site p-values from the Cox proportional hazards model adjusted for age, sex, smoking status, education level, BMI as a continuous variable, and the first 10 genetic principal components (this model fails the PH assumption). Technical covariates (chip ID, chip row, study center, visit, project) and cell type proportions were adjusted too for both models. Note, not using BMI x time interaction failed the PH assumption in Black adults alone; consequently there was noticeable variation in the CpG site p-values from Black adults (and not from White adults), which was also reflected in the 'All' (meta-analyzed) group.

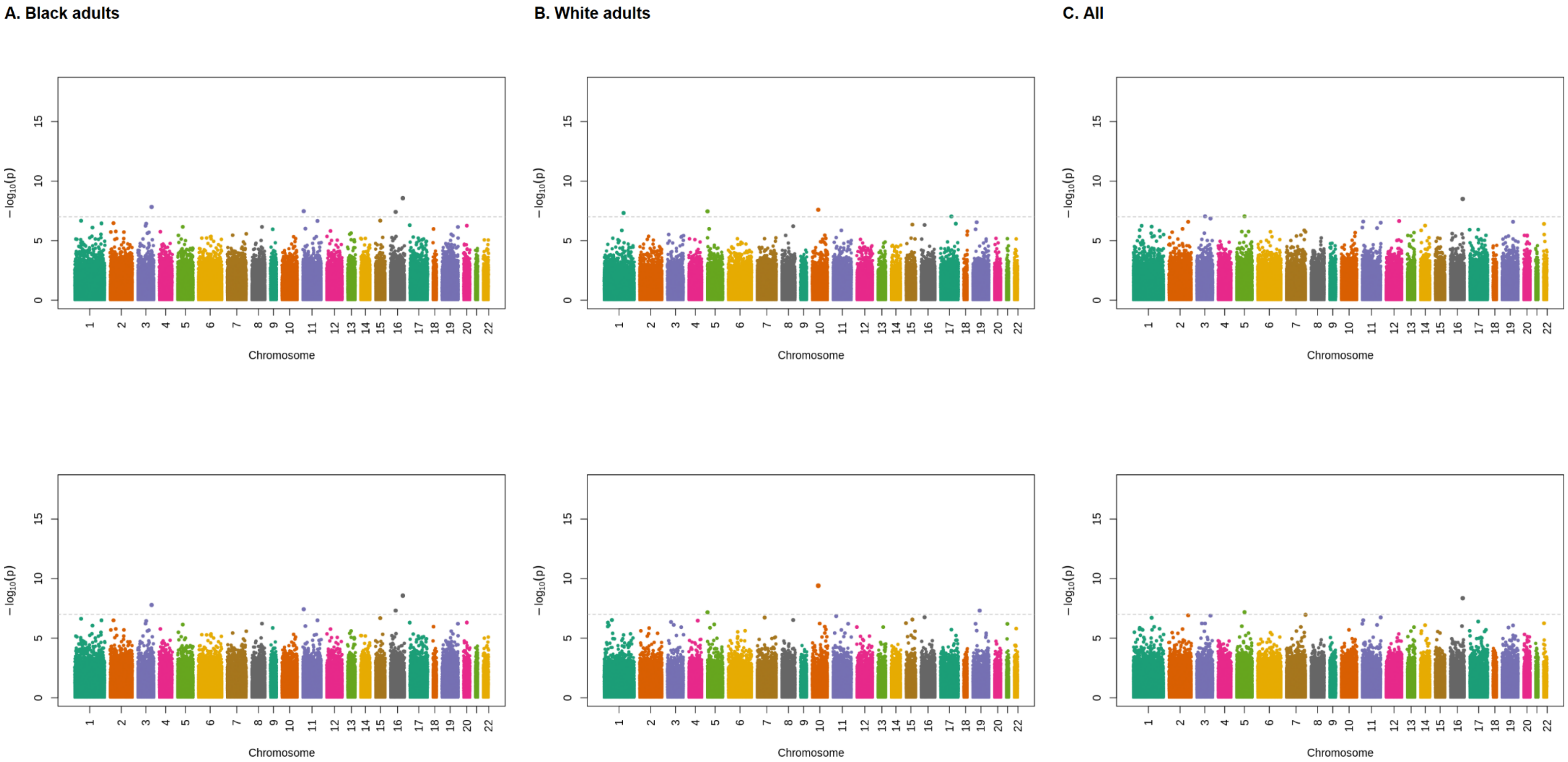


**Supplementary Figure 16**: Manhattan plots for fasting glucose-adjusted Cox proportional hazards models showing differences in DNAm associations with and without violation of proportional hazards (PH) assumptions. For each race group, the top Manhattan plot shows the CpG site p-values from the Cox proportional hazards model adjusted for age, sex, smoking status, education level, fasting glucose with time-varying effects across periods of time since time origin, and the first 10 genetic principal components (this model satisfies the PH assumption). The bottom Manhattan plot shows the CpG site p-values from the Cox proportional hazards model adjusted for age, sex, smoking status, education level, fasting glucose as a continuous variable, and the first 10 genetic principal components (this model fails the PH assumption). Note, technical covariates (chip ID, chip row, study center, visit, project) and cell type proportions were adjusted too for both models.

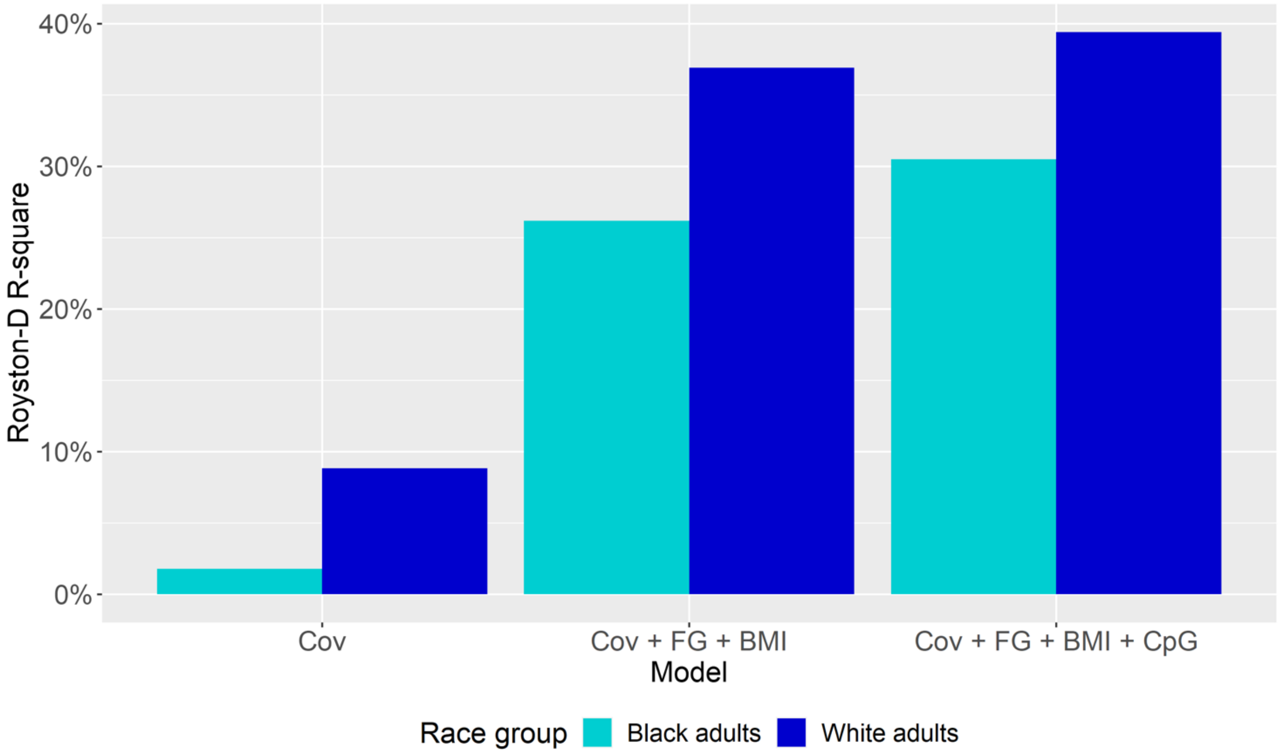


**Supplementary Figure 17:** Percent variance explained in incident type 2 diabetes in Black and White adults separately across 3 nested Cox proportional hazards models. Model "Cov" includes covariates age, sex, smoking status, education level and the first 10 genetic principal components. Model "Cov + FG + BMI" includes covariates age, sex, smoking status, education level, BMI, fasting glucose, and the first 10 genetic principal components. Model "Cov + FG + BMI + CpG" additionally includes significantly associated CpG sites discovered in our epigenome-wide incident diabetes analysis pertaining to those race groups (8 sites for Black adults and 2 sites for White adults).

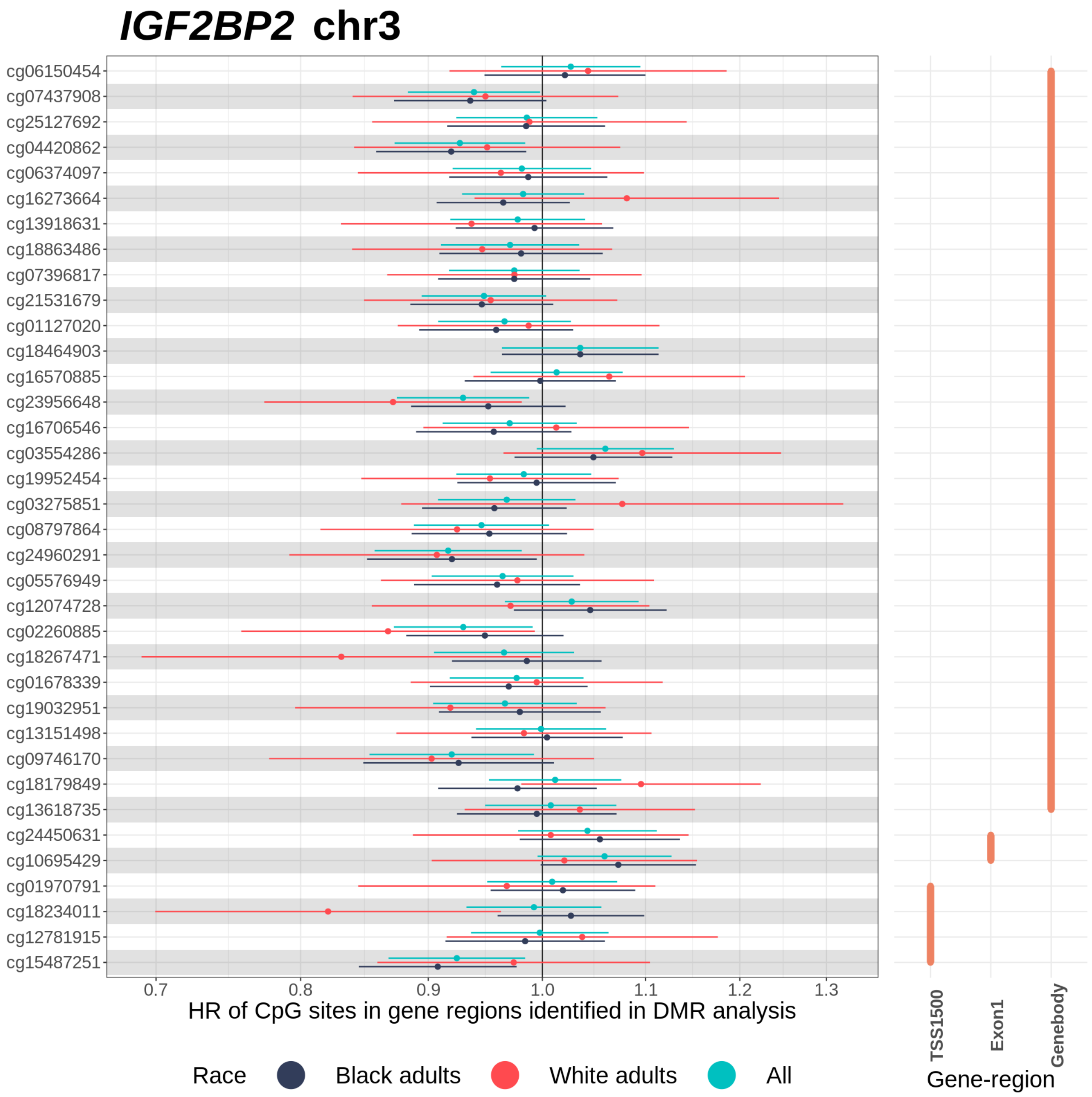


**Supplementary Figure 18:** Effect size plot of CpG sites annotated to gene-regions of *IGF2BP2* on chromosome 3. The graph on the left depicts hazard ratio estimates of CpG sites with 95% confidence intervals across race groups obtained from Cox proportional hazards model adjusting for age, sex, smoking status, education level, and the first 10 genetic principal components. CpG sites are arranged from top to bottom in ascending order of chromosomal position. The graph on the right indicates the gene-region DMR groups that each CpG site was included in.

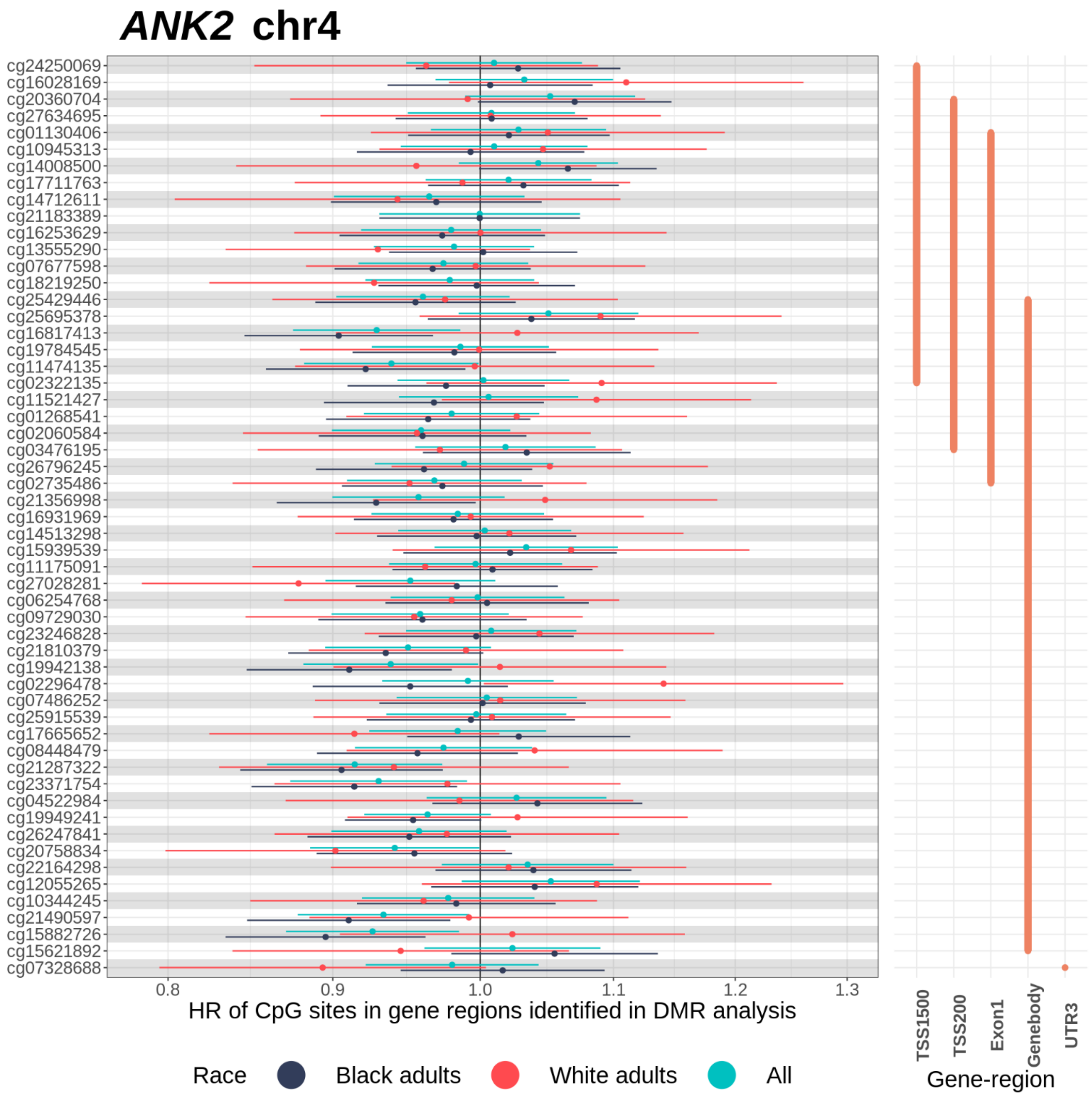


**Supplementary Figure 19:** Effect size plot of CpG sites annotated to gene-regions of *ANK2* on chromosome 4. The graph on the left depicts hazard ratio estimates of CpG sites with 95% confidence intervals across race groups obtained from Cox proportional hazards model adjusting for age, sex, smoking status, education level, and the first 10 genetic principal components. Technical covariates (chip ID, chip row, study center, visit, project) and cell type proportions were adjusted too. CpG sites are arranged from top to bottom in ascending order of chromosomal position. The graph on the right indicates the gene-region DMR groups that each CpG site was included in.

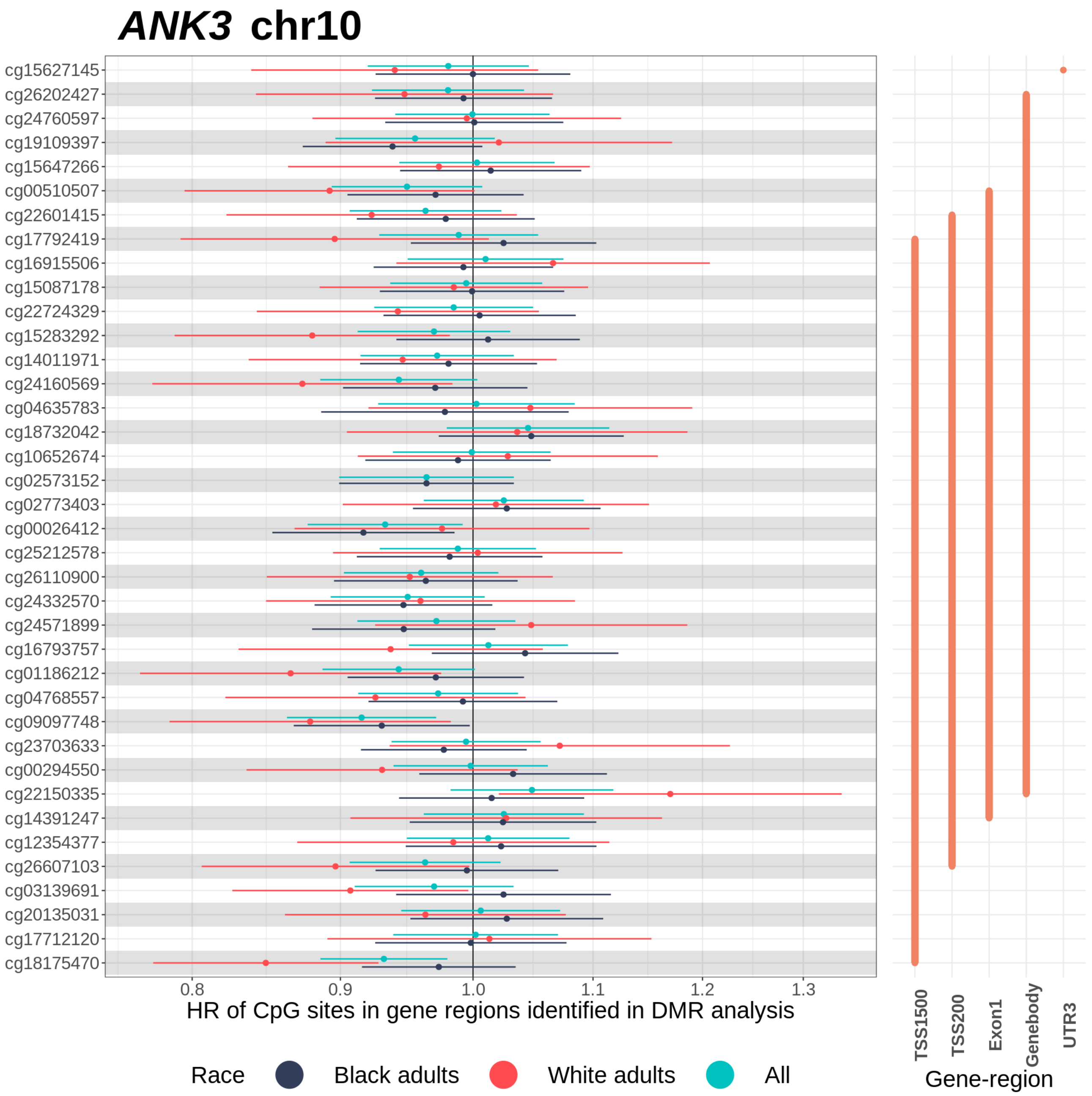


**Supplementary Figure 20:** Effect size plot of CpG sites annotated to gene-regions of *ANK3* on chromosome 10. The graph on the left depicts hazard ratio estimates of CpG sites with 95% confidence intervals across race groups obtained from Cox proportional hazards model adjusting for age, sex, smoking status, education level, and the first 10 genetic principal components. Technical covariates (chip ID, chip row, study center, visit, project) and cell type proportions were adjusted too. CpG sites are arranged from top to bottom in ascending order of chromosomal position. The graph on the right indicates the gene-region DMR groups that each CpG site was included in.

**A**

**chr5:110062343-110062837**

| Position | CpG | Blacks HR (95% CI) | Whites HR (95% CI) | All HR (95% CI) |
|---|---|---|---|---|
| 110062343 | cg23279021 | 0.92 (0.86, 0.99) | 0.86 (0.75, 0.98) | 0.91 (0.85, 0.96) |
| 110062384 | cg17248924 | 0.92 (0.86, 0.99) | 0.84 (0.74, 0.96) | 0.90 (0.85, 0.96) |
| 110062398 | cg11641395 | 0.91 (0.85, 0.98) | 0.88 (0.77, 1.00) | 0.90 (0.85, 0.96) |
| 110062417 | cg06429214 | 0.90 (0.84, 0.97) | 0.91 (0.80, 1.04) | 0.91 (0.85, 0.97) |
| 110062473 | cg25259944 | 0.91 (0.84, 0.97) | 0.85 (0.75, 0.97) | 0.89 (0.84, 0.95) |
| 110062729 | cg19526166 | 0.91 (0.84, 0.97) | 0.88 (0.78, 1.00) | 0.90 (0.85, 0.96) |
| 110062837 | cg06414816 | 0.91 (0.85, 0.97) | 0.89 (0.79, 1.01) | 0.90 (0.85, 0.96) |

Blacks
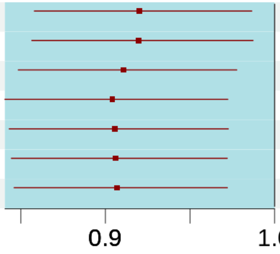


Whites
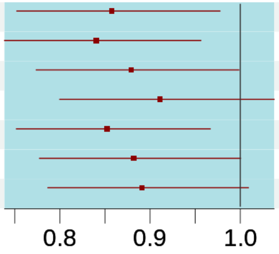


All
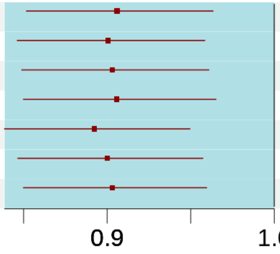


**B**

**chr16:50321678-50322156**

| Position | CpG | Blacks HR (95% CI) | Whites HR (95% CI) | All HR (95% CI) |
|---|---|---|---|---|
| 50321678 | cg13851211 | 1.07 (0.99, 1.15) | 1.11 (0.97, 1.26) | 1.08 (1.01, 1.15) |
| 50321818 | cg02879453 | 1.11 (1.03, 1.21) | 1.29 (1.13, 1.48) | 1.16 (1.08, 1.24) |
| 50321878 | cg08021532 | 1.09 (1.00, 1.18) | 1.11 (0.95, 1.29) | 1.09 (1.02, 1.17) |
| 50322074 | cg06897661 | 1.16 (1.07, 1.25) | 1.21 (1.05, 1.39) | 1.17 (1.09, 1.25) |
| 50322156 | cg23580000 | 1.15 (1.06, 1.24) | 1.17 (1.04, 1.32) | 1.16 (1.08, 1.23) |

Blacks
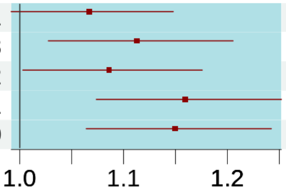


Whites
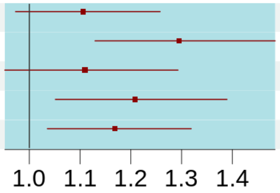


All
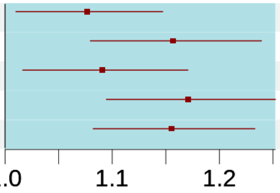


**C**

**chr6:33040535-33041697**

| Position | CpG | Blacks HR (95% CI) | Whites HR (95% CI) | All HR (95% CI) |
|---|---|---|---|---|
| 33040535 | cg25824217 | 1.05 (0.97, 1.13) | 0.88 (0.77, 1.00) | 1.00 (0.94, 1.07) |
| 33040610 | cg20617328 | 1.07 (1.00, 1.15) | 0.91 (0.80, 1.03) | 1.03 (0.97, 1.10) |
| 33040769 | cg00403478 | 1.08 (1.00, 1.16) | 0.85 (0.75, 0.97) | 1.02 (0.96, 1.09) |
| 33040914 | cg13906813 | 1.11 (1.03, 1.19) | 0.79 (0.70, 0.90) | 1.02 (0.96, 1.09) |
| 33041218 | cg15575880 | 1.07 (1.00, 1.15) | 1.00 (0.88, 1.13) | 1.05 (0.99, 1.12) |
| 33041221 | cg20907136 | 1.07 (1.00, 1.15) | 1.02 (0.90, 1.16) | 1.06 (1.00, 1.13) |
| 33041229 | cg01804934 | 1.07 (0.99, 1.15) | 0.88 (0.77, 1.00) | 1.02 (0.96, 1.09) |
| 33041268 | cg24287218 | 1.08 (1.00, 1.16) | 0.82 (0.72, 0.94) | 1.01 (0.95, 1.08) |
| 33041343 | cg09321817 | 1.09 (1.01, 1.18) | 0.88 (0.77, 1.00) | 1.03 (0.97, 1.10) |
| 33041406 | cg13031167 | 1.08 (1.01, 1.16) | 0.86 (0.75, 0.98) | 1.03 (0.96, 1.09) |
| 33041583 | cg11451043 | 1.09 (1.02, 1.17) | 0.94 (0.83, 1.08) | 1.06 (0.99, 1.12) |
| 33041697 | cg00798281 | 1.08 (1.01, 1.16) | 1.03 (0.91, 1.16) | 1.07 (1.00, 1.13) |

Blacks
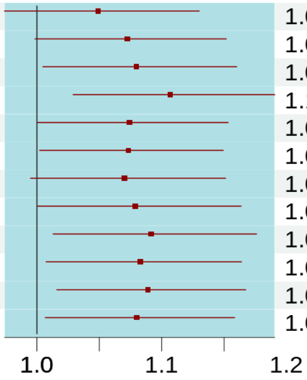


Whites
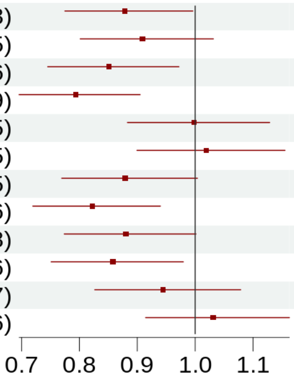


All
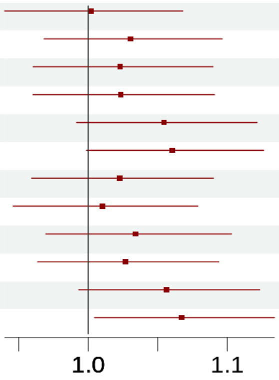

D

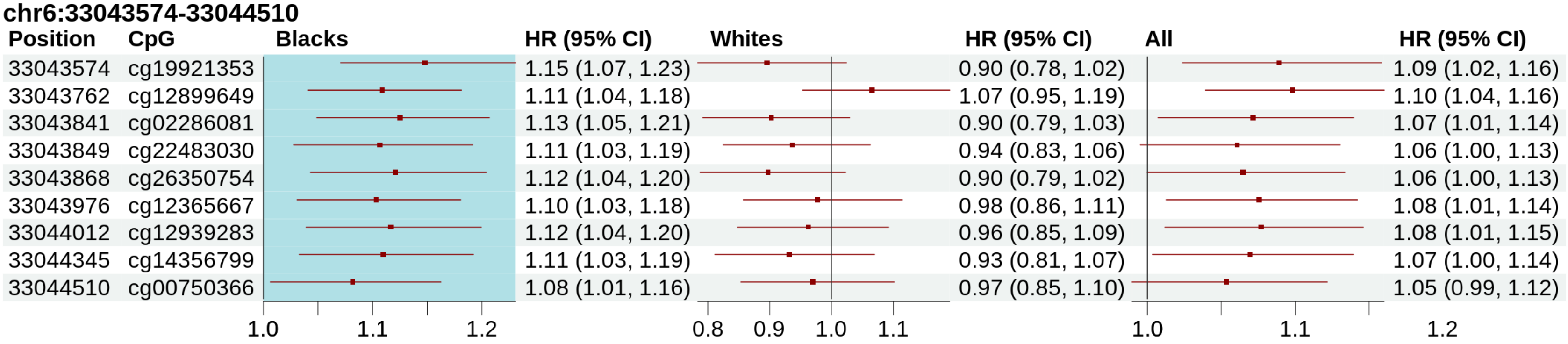
chr6:33043574-33044510
Position CpG Blacks HR (95% CI) Whites HR (95% CI) All HR (95% CI)
33043574 cg19921353 1.15 (1.07, 1.23) 0.90 (0.78, 1.02) 1.09 (1.02, 1.16)
33043762 cg12899649 1.11 (1.04, 1.18) 1.07 (0.95, 1.19) 1.10 (1.04, 1.16)
33043841 cg02286081 1.13 (1.05, 1.21) 0.90 (0.79, 1.03) 1.07 (1.01, 1.14)
33043849 cg22483030 1.11 (1.03, 1.19) 0.94 (0.83, 1.06) 1.06 (1.00, 1.13)
33043868 cg26350754 1.12 (1.04, 1.20) 0.90 (0.79, 1.02) 1.06 (1.00, 1.13)
33043976 cg12365667 1.10 (1.03, 1.18) 0.98 (0.86, 1.11) 1.08 (1.01, 1.14)
33044012 cg12939283 1.12 (1.04, 1.20) 0.96 (0.85, 1.09) 1.08 (1.01, 1.15)
33044345 cg14356799 1.11 (1.03, 1.19) 0.93 (0.81, 1.07) 1.07 (1.00, 1.14)
33044510 cg00750366 1.08 (1.01, 1.16) 0.97 (0.85, 1.10) 1.05 (0.99, 1.12)
1.0 1.1 1.2
0.8 0.9 1.0 1.1
1.0 1.1 1.2

E

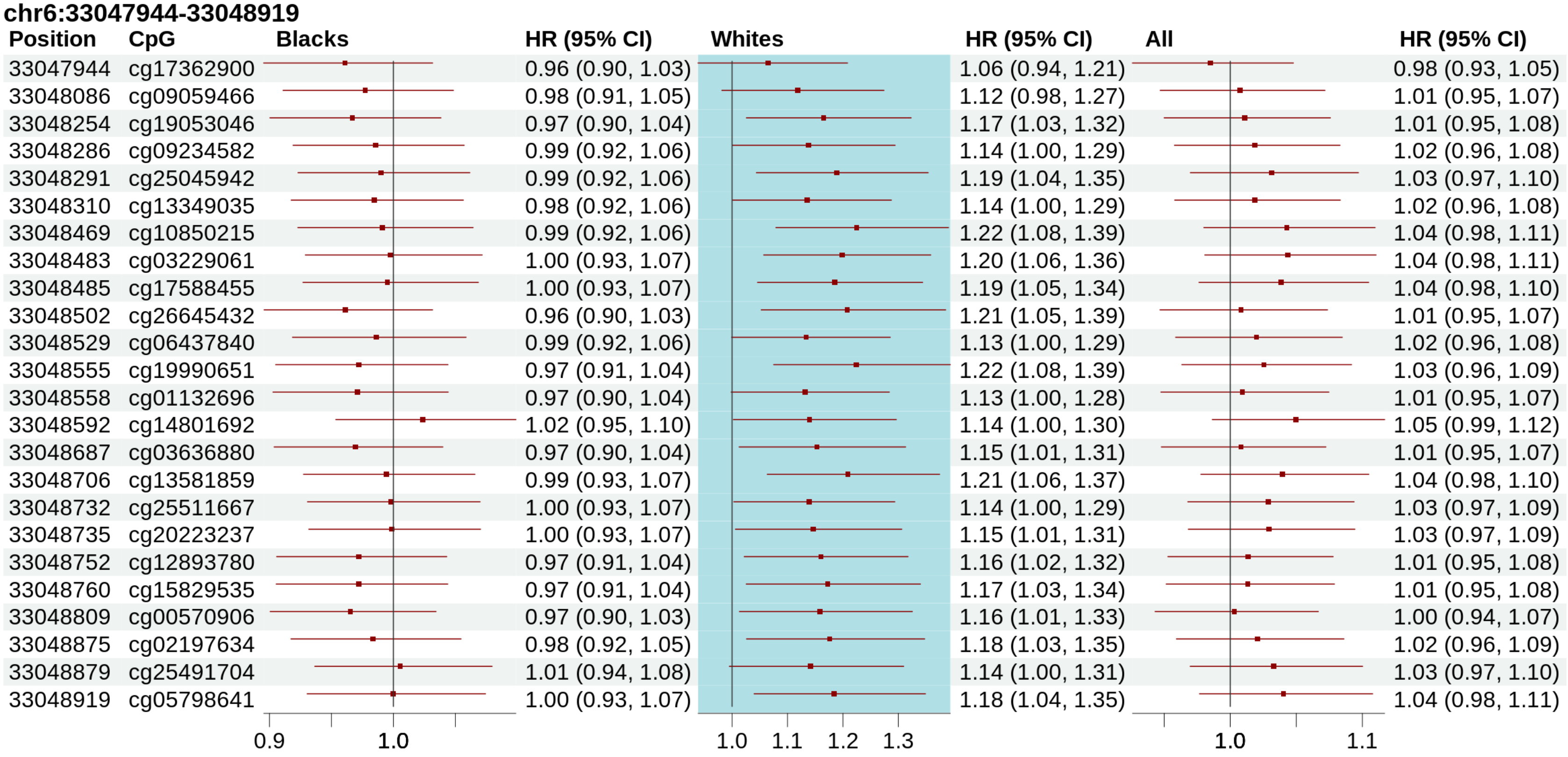
chr6:33047944-33048919
Position CpG Blacks HR (95% CI) Whites HR (95% CI) All HR (95% CI)
33047944 cg17362900 0.96 (0.90, 1.03) 1.06 (0.94, 1.21) 0.98 (0.93, 1.05)
33048086 cg09059466 0.98 (0.91, 1.05) 1.12 (0.98, 1.27) 1.01 (0.95, 1.07)
33048254 cg19053046 0.97 (0.90, 1.04) 1.17 (1.03, 1.32) 1.01 (0.95, 1.08)
33048286 cg09234582 0.99 (0.92, 1.06) 1.14 (1.00, 1.29) 1.02 (0.96, 1.08)
33048291 cg25045942 0.99 (0.92, 1.06) 1.19 (1.04, 1.35) 1.03 (0.97, 1.10)
33048310 cg13349035 0.98 (0.92, 1.06) 1.14 (1.00, 1.29) 1.02 (0.96, 1.08)
33048469 cg10850215 0.99 (0.92, 1.06) 1.22 (1.08, 1.39) 1.04 (0.98, 1.11)
33048483 cg03229061 1.00 (0.93, 1.07) 1.20 (1.06, 1.36) 1.04 (0.98, 1.11)
33048485 cg17588455 1.00 (0.93, 1.07) 1.19 (1.05, 1.34) 1.04 (0.98, 1.10)
33048502 cg26645432 0.96 (0.90, 1.03) 1.21 (1.05, 1.39) 1.01 (0.95, 1.07)
33048529 cg06437840 0.99 (0.92, 1.06) 1.13 (1.00, 1.29) 1.02 (0.96, 1.08)
33048555 cg19990651 0.97 (0.91, 1.04) 1.22 (1.08, 1.39) 1.03 (0.96, 1.09)
33048558 cg01132696 0.97 (0.90, 1.04) 1.13 (1.00, 1.28) 1.01 (0.95, 1.07)
33048592 cg14801692 1.02 (0.95, 1.10) 1.14 (1.00, 1.30) 1.05 (0.99, 1.12)
33048687 cg03636880 0.97 (0.90, 1.04) 1.15 (1.01, 1.31) 1.01 (0.95, 1.07)
33048706 cg13581859 0.99 (0.93, 1.07) 1.21 (1.06, 1.37) 1.04 (0.98, 1.10)
33048732 cg25511667 1.00 (0.93, 1.07) 1.14 (1.00, 1.29) 1.03 (0.97, 1.09)
33048735 cg20223237 1.00 (0.93, 1.07) 1.15 (1.01, 1.31) 1.03 (0.97, 1.09)
33048752 cg12893780 0.97 (0.91, 1.04) 1.16 (1.02, 1.32) 1.01 (0.95, 1.08)
33048760 cg15829535 0.97 (0.91, 1.04) 1.17 (1.03, 1.34) 1.01 (0.95, 1.08)
33048809 cg00570906 0.97 (0.90, 1.03) 1.16 (1.01, 1.33) 1.00 (0.94, 1.07)
33048875 cg02197634 0.98 (0.92, 1.05) 1.18 (1.03, 1.35) 1.02 (0.96, 1.09)
33048879 cg25491704 1.01 (0.94, 1.08) 1.14 (1.00, 1.31) 1.03 (0.97, 1.10)
33048919 cg05798641 1.00 (0.93, 1.07) 1.18 (1.04, 1.35) 1.04 (0.98, 1.11)
0.9 1.0
1.0 1.1 1.2 1.3
1.0 1.1

F

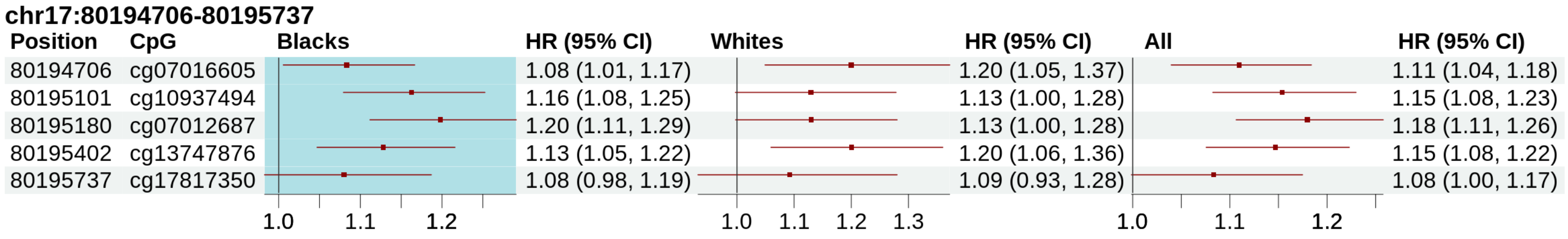


G

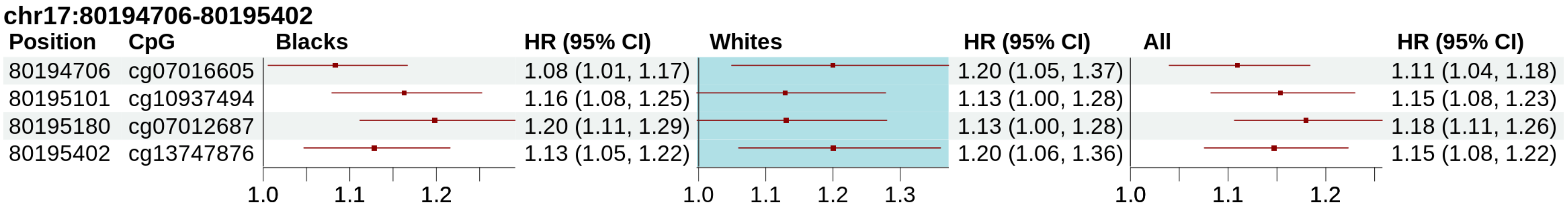


**Supplementary Figure 21**: Effect size plots of individual CpG sites in each differentially methylated region (DMR) across race groups. Columns shaded in blue represent race groups in which the corresponding DMR was significantly associated with incident type 2 diabetes. Group-specific DMRs were identified using Šidák-corrected significance threshold of 5%. DMR analysis of each race group was performed using comb-p on p-values of CpG site associations from the Cox proportional hazards model adjusting for age, sex, smoking status, education level and first 10 genetic principal components. Technical covariates (chip ID, chip row, study center, visit, project) and cell type proportions were adjusted too.

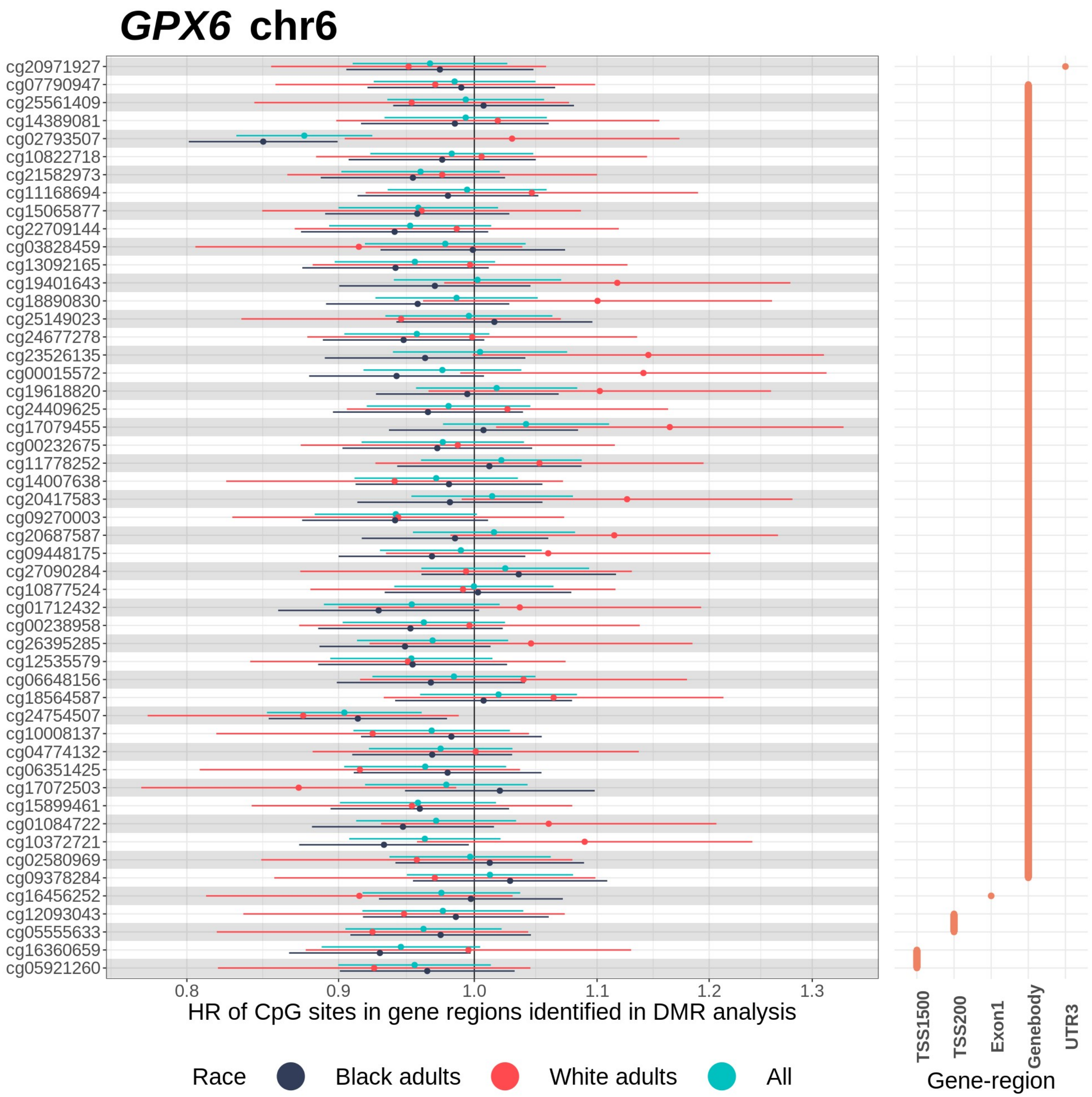


**Supplementary Figure 22:** Effect size plot of CpG sites annotated to gene-regions of *GPX6* on chromosome 6. The graph on the left depicts hazard ratio estimates of CpG sites with 95% confidence intervals across race groups obtained from Cox proportional hazards model adjusting for age, sex, smoking status, education level, and the first 10 genetic principal components. Technical covariates (chip ID, chip row, study center, visit, project) and cell type proportions were adjusted too. CpG sites are arranged from top to bottom in ascending order of chromosomal position. The graph on the right indicates the gene-region DMR groups that each CpG site was included in.
